# Do You Know Your Daily Antibiotic Intake through Residues in Your Diet?

**DOI:** 10.1101/2023.12.01.23299305

**Authors:** Jegak Seo, Frank Kloprogge, Andrew M. Smith, Kersti Karu, Lena Ciric

**Author notes:** Corresponding author: Jegak Seo, Healthy Infrastructure Research Group, Department of Civil, Environmental and Geomatic Engineering, University College London, Gower Street, London WC1E 6BT.

## Abstract

While the use of a wide range of antibiotics has been reported as extensive in the rearing of agricultural animals, extremely limited information is available on the antibiotic residues in animal products and the adverse impact consistent low-level exposure to antibiotics might have on the human body as well as its microbiome. The aim of this study was to estimate the possible antibiotic concentrations humans are exposed to via their diet using the concentration of antibiotics in animal food products and water, and an online survey on dietary habits. A total of 131 participants completed the dietary habits survey, the majority belonging to the omnivorous diet habit. Distinct dietary trends were observed into omnivorous and unknown groups eating food-producing animal products, with specific food types dominating each meal: pork (e.g. ham) and dairy products (e.g. milk, yoghurt) during breakfast, beef (e.g. burger) and chicken (e.g. chicken breast) products during lunch, and fish (e.g. salmon fillet) during dinner. 34 different animal-based food and drink products were tested for the presence of ten different antibiotics. Low levels of nine of the ten antibiotics were detected across the samples tested with amoxicillin and trimethoprim being the most frequently detected antibiotics from all samples with concentrations ranging from 216.7-6866.9 μg/kg and 55.2-461.7 μg/kg, respectively. Of all products tested, over 35% exceeded the acceptable daily intake antibiotic concentration for amoxicillin, ampicillin, and enrofloxacin.

## 1. Introduction

Antimicrobial resistance (AMR) poses a significant global threat and has the potential to become the next pandemic. Currently, AMR is responsible for 700,000 deaths annually, and without substantial policy changes, this number is projected to escalate to 10 million deaths per year by 2050, as highlighted by the report from Jim O’Neil in 2016 (O’Neill, 2016). Extensive research efforts have primarily concentrated on the direct intake of antibiotics by humans through prescriptions, pharmacy purchases, and hospital use (Ayukekbong, Ntemgwa, & Atabe, 2017; Llor & Bjerrum, 2014; van Staa et al., 2020). However, it is becoming increasingly evident that exposure risks associated with the consumption of food-producing animals, including meats and dairy products, are of growing concern. Excessive agricultural and veterinary antibiotic usage has led to the pervasive detection of veterinary antibiotic residues in animal products worldwide, creating a vicious cycle of escalating veterinary antibiotic use driven to the increasing levels of antibiotic resistance in animals (Andrew Bamidele & Oluwakamisi Festus, 2019; Economou & Gousia, 2015; FAO, 2016; Treiber & Beranek-Knauer, 2021). Furthermore, the control and regulation of veterinary antibiotic purchase and usage pose significant challenges in most countries (Manyi-Loh, Mamphweli, Meyer, & Okoh, 2018), resulting in the inappropriate use of antibiotics in animals as growth promoters and for treatment without adhering to prescribed withdrawal periods (Morehead & Scarbrough, 2018).

Antibiotics have been detected in food products and drinking water due to a wide range of antibiotic used not only for treatment in infectious disease but also in agricultural run-off, wastewater treatment, non-medical applications, and open defecations (Manyi-Loh et al., 2018). The identification and quantification of antibiotics in food and drinking water has become a new field of study to exploring the undiscovered world which is harmfully polluted with variety of antibiotics by humans since 2000s (Kraemer, Ramachandran, & Perron, 2019). A recent comprehensive review of antibiotic monitoring studies conducted throughout the world identified residues of antibiotics which are used in humans and animals in meat and dairy products, plants and drinking water (Klein et al., 2018; Manyi-Loh et al., 2018). Antibiotics administered to humans are frequently detected in food and drinking water, and their presence is also often observed in plants, likely due to exposure through irrigation or the use of fertilizers derived from wastewater and manure (Ali Mirza et al., 2020; Stockwell & Duffy, 2012). These studies recommend that the risk of AMR through chronic consumption of a trace level of antibiotics in foods or drinks is significant.

The overall aim of this study was to estimate daily intake of antibiotic residues via diet habits using antibiotic concentrations present in drinking water and animal-based food products from the UK establishing a measure of the subsequent risk of human exposure. Specifically, this research aimed to explore the antibiotic concentration in food products, including beef, pork, chicken and fish, dairy products, and drinking water by monitoring questionnaires and analysis of food samples collected in local stores. Therefore, this study was delivered through three specific objectives: (1) to characterise, using an online questionnaire, how public populations can come into contact with antibiotics through the consumption of food-producing animal products and drinking water, (2) to determine the levels of antibiotic concentrations in food-producing animal products and drinking water by collecting samples from large supermarket chains, (3) to analyse the range of antibiotic exposure among survey participants by combining data on their dietary habits with the concentration of antibiotic residues in food-producing animal products an drinking water.

## 2. Materials and Methods

### 2.1. Online diet survey

The online survey is used as a direct method for dietary assessment which collects primary dietary data from individuals (FAO, 2018). UCL Opinio (https://opinio.ucl.ac.uk/admin/folder.do) was used to apply a quantitative method to determine both types and amounts of food consumed. Ethical approval for the questionnaire was provided by the UCL Research Ethics Committee, project number 19139/001.

Two days of 24-hour recall and food frequency questionnaire (FFQ) were the main channel using a retrospective approach; estimated food record and weighed food record were included as subsidiary functions using Likert scales and open-ended questions; and innovative technologies supported by any devices, were used to support the technical approaches for the participants and increasing the accuracy of the survey. In the 24-hour recall section, participants were asked to recall the intakes for 48 hours in total. Twenty slots for food or drinks per day were provided to ensure sufficient opportunities to record all animal-based food and drinks consumed. Time, place, type and name of product, and volume (ml) or mass (g) of product were required for each different food or drink type. After 24-hour recall, FFQ was assessed to investigate the frequency with which foods and drinks, and/or food groups were consumed over a certain time period. After completing the two sections, the participants were asked to compare their dietary history to their general intake in a week using Likert scales. Firstly, participants were asked to estimate the number of intake days per week. Then, the amount of each recorded food and drink was compared to the general intake in a week by rating in percentage, on a Likert scale of between less than 10% or more than 200%.

In accordance with the FAO guidelines (2018), the survey was designed to facilitate a nuanced analysis of results. The focus was placed on the detailed collection of dietary histories, which was pivotal for the estimation of antibiotic consumption. The US FDA’s Estimated Meal Intake formula (equation 1), which standardises the weight assumption for an adult participant at 60 kg, was utilized for this purpose (FDA, 2018). In this research, the reference to 12 o’clock was intended to encompass the time range between 1200 and 1259, and similarly, other hourly references were aligned with their respective one-hour time intervals.

### 2.2. Antibiotic quantification in food and drink

#### 2.2.1. Antibiotics, chemicals and reagents

The following antibiotics (CAS number): tetracycline (64-75-5), oxytetracycline (6153-64-6), amoxicillin (61336-70-7), ampicillin (7177-48-2), trimethoprim (738-70-5), sulfadiazine (68-35-9), ciprofloxacin (85721-33-1), enrofloxacin (93106-60-6), erythromycin (114-07-8), and tylosin (1405-54-5), were purchased from Sigma-Aldrich (St. Louis, MO, USA), all with purities ≥ 99%. All reagents chromatographic grade acetonitrile (ACN), water, formic acid (FA) with purity higher than 99.8% used for LC-MS analysis were ordered from Fisher Scientific (Lancashire, UK).

#### 2.2.2. Low Temperature Partitioning Extraction (LTPE)

All samples were purchased from large supermarket chains in London, United Kingdom. The samples were purchased at the same time as when the survey was open to participants, i.e. 28/May/2021 to 30/Jul/2021 and 12/Jan/2022 to 17/Mar/2022.

For all sample preparation, at least of 3.0 g of a whole food or drink sample was homogenized using a kitchen blender (BOSCH, MSM6B150GB) for 1 min in triplicate. 1.0 g of the homogenized sample was aliquoted to a 50 mL test tube. Aliquoted replicates were further homogenized using pellet pestles (Bel-ART SP SCIENCEWARE, 19923-000) for 1 min. The processed sample vials were covered with aluminium foil and stored at -20 C prior to analysis. 1.0 g of HPLC-LiChropur™ NaCl (Merck, 7647-14-5) was added to the tube and vortexed at 448 xg for 1 min, followed by addition of 8.0 mL of Acetonitrile 50%, water 47.5%, TFA 2.5% (Honeywell, 19182-250mL) to the tube. The tube was vortexed and centrifuged for another 5 mins. The prepared samples were stored at -20 C freezer overnight. Then, 1.5 mL of the organic phase was removed and transferred to a 2.0 mL microcentrifuge tube. The samples were centrifuged at 3,278 xg for 10 mins at 25 C and 1.0 mL of supernatant was transferred to individual HPLC glass vial for LC-MS analysis.

#### 2.2.3. LC-MS analysis

Samples were analysed using a liquid chromatography tandem mass spectrometry (LC-MS/MS) instrument. The instrument consisted of an Accela LC system connected to a Finnigan Linear Trap Quadrupole (LTQ) Linear Ion Trap mass spectrometer from Thermo Fisher Scientific, UK. The chromatographic separation was achieved using a Hypersil GOLD C18 column (150 mm x 2.1 mm, 1.9 μm; Thermo Fisher Scientific, UK). The column temperature was maintained at 30 C. Mobile phases A and B were: (A) water with 0.1% formic acid and (B) acetonitrile with 0.1% formic acid, and the flow rate was 200 μL/min. The gradient program was as follows: 2% of B for the first 2 min and after gradual change to 98% B in 16 minutes and changed to 2% of B in 0.1 min and remained at 2% B for another 1.9 minutes. The total run time was 20 minutes per sample. The injected sample volume was 10 μL. The liquid effluent from the C18 column was directed into the electrospray (ESI) source of the LTQ mass spectrometer (MS). The ESI was in positive mode and the source parameters were as follows: a spray voltage of 4500 V, capillary temperature set to 280 °C, sheath gas at a pressure of 40 psi, ion sweep gas pressure (0 psi), auxiliary gas set at 5 psi, and a skimmer offset at 25 V. The data was collected using a full-scan MS event with a mass range from m/z 50 to 1000 and in the MS/MS event, which was set-up for each m/z value antibiotic corresponding to each antibiotic as per Figure S2. The isolation width was 2.0 and a collision energy of 35. The analytical batch was set-up containing water blanks (H_2_O), which were analysed after each sample analysis and a quality control which consisted of a pure antibiotic at concentration of 10 µg/L.

#### 2.2.4. The method validation

Figure S2 shows chromatographic separation of 10 antibiotics on the C18 column, and their retention times are summarised in Table S3. The LC-MS method validation parameters such as accuracy, limit of detection (LOD), and limit of quantification (LOQ) were calculated and summarised in Supplementary Materials Table S1. For calibration curves, five replicates at nominal concentrations of 50, 100, and 500 μg/L were prepared and analysed by LC-MS. An accuracy (%) and relative standard deviation (RSD; %) of the measurements were determined and the calibration curves were constructed for each antibiotic (Table S3). The accuracy and RSD ranged between 97.2 to 111.22% and 0.01 to 0.92%, respectively.

The LTPE validation is summarised in Supplementary Materials. Pork chop meat was used as a pure matrix to compare the accuracy of the LTPE methods. Triplicates of the non-spiked pure matrix were tested with LTPE methods to determine the presence of antibiotics. Triplicates of the pure matrix were spiked at the nominal concentrations of 100 μg/L of 10 antibiotic mixture solution and the antibiotics were extracted with the LTPE methods. Linear regression analysis was carried to calculate the linearity (R >0.999) of the calibration curves by Microsoft Excel version 16.53 (Microsoft Excel, 2021) and results are summarised in Table S4. The recovery of LTPE method using 100 ug/kg of 10 antibiotic mixture stock solution was between 87.6 to 93.5%, and the recovery of using triplicates of pork chop matrix spiked with a 100 μg/kg of 10 antibiotics mixture was between 89.6 to 95.4% (Table S4).

### 2.3. Estimated Meal Intake (EMI)

The estimated daily intake formula from the US FDA (FDA, 2018) was modified to calculate the antibiotic intake from each meal instead of the total intake of substances in a day by equation 1. Also, additional dilution factors such as average volume of drinks and meal, stomach acid and bile juice in the human digestive system were taken account to determine the luminal concentration of antibiotic in human duodenum.

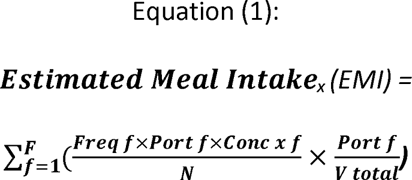

Where,

F = Total number of foods in which antibiotic “x” can be found
Freq_f_ = Average portion size for food “f”
Port_f_ = Number of eating occasions of food “f” over “N” meals in during the survey
Conc_xf_ = Concentration of the antibiotic “x” in food “f”
N = Total number of meals in the survey
V_total_ = Average volume of drinks + average volume of meal + average volume of human stomach juice (60 ml)

The most frequently consumed meat type was chosen for the representative food type of each meal. The list of consumed foods and drinks were determined in each meal over the 48hr diet survey. The detected antibiotics were determined from the specified foods and drinks and the average concentrations of detected antibiotics were applied. However, any concentrations below the ADI concentration (https://apps.who.int/food-additives-contaminants-jecfa-database/) were excluded to the list for each food item. The total volume of each meal was calculated by adding the average volume of the consumed drinks and foods during a meal and average volume of gut juice, 60 ml, which is the volume when the human stomach is empty (Schiller et al., 2005).

### 2.5. Statistical Analysis

Regression analysis was carried to determine the accuracy and validity (R >0.999) of calibration curves for the antibiotic measurement using LC-MS. The mean difference of food and drink consumption in different days and seasons were statistically analysed and compared by one-way ANOVA test with post-hoc Bonferroni test.

## 3. Results and Discussion

### 3.1. Demographical profiles and overall consumption trend

The online survey to investigate participants’ dietary habits over 48 hours was conducted between 28/05/2021 and 30/07/2021 in summer (n= 51) and 12/01/2022 and 17/03/2022 (n= 80; dd/mm/yyyy). All participants (n= 131) agreed to the UCL General Research Participant Privacy Notice.

The total participant count for this research was adjusted to 117 (45 from the summer survey and 72 from the winter survey), encompassing both omnivores and individuals with unknown dietary habits as it provided a diverse representation of food-producing animal product dietary habits and a statistically sufficient number of respondents (FAO, 2018; UK Government, 2021).

**Table 1.** Profile of participants in the summer and winter dietary habit survey (2021/2022).

| Season (Year) | Participant Age |  |  |  |  |  |
| --- | --- | --- | --- | --- | --- | --- |
|  | 20s<br>(18-29) | 30s<br>(30-39) | 40s<br>(40-49) | 50s<br>(50-59) | 60s<br>(60-69) | 70s<br>(>70) |
|  | *n* | *n* | *n* | *n* | *n* | *n* |
| Summer (2021)<sup>a</sup> | 39 | 13 | 8 | 0 | 1 | 0 |
| Winter (2022)<sup>b</sup> | 33 | 17 | 15 | 11 | 3 | 1 |
| Total<br>*n* = 141 (21/22) | 62 | 30 | 23 | 11 | 4 | 1 |

| Season (Year) | Dietary Habits |  |  |  |  |  |
| --- | --- | --- | --- | --- | --- | --- |
|  | Omnivorous | Vegetarian | Vegan | Protein-based | Halal | Unknown |
|  | *n* | *n* | *n* | *n* | *n* | *n* |
| Summer (2021)<sup>a</sup> | 30 | 3 | 2 | 1 | 0 | 15 |
| Winter (2022)<sup>b</sup> | 70 | 6 | 0 | 0 | 2 | 2 |
| Total<br>*n* = 141 (21/22) | 100 | 9 | 2 | 1 | 2 | 17 |

<sup>a</sup> Summer data was collected from 28/05/2021 to 30/07/2021.
<sup>b</sup> Winter data was collected from 12/01/2022 to 17/03/2022.

To capitulate briefly, we have shown that our survey results are in line with previously published research and national surveys in the UK (UK Government, 2021). In general, meat consumption followed the peaks of water consumption. In both seasons, only pork was consumed during breakfast over the two days. Similarly, chicken and beef were mostly consumed at lunch. Fish and chicken were the most frequently consumed animal products, respectively, in both seasons. Most of the participants had meals at regular times without special occasions such as celebrations or irregular skipping meals.

UK adults in the national survey (n= 8174) reported consuming pork products such as ham, bacon, and sausages the most at breakfast(Gaal, Kerr, Ward, McNulty, & Livingstone, 2018). Moreover, the participants had the highest consumption of beef > chicken > fish over the rest of the day. It was also determined from the national survey that the most consumed meats in the UK were (in order of highest to lowest) beef, chicken, and fish(Gaal et al., 2018). It is reasonable to assume that the participants dairy products intake is most via drinking milk at breakfast. Water intake is directly related to food consumption.

### 3.2. Meat consumption

Based on the overall meat consumption trend, the peaks of meat consumptions are shown in figure 1. In summer and winter, both had the same pattern of food product type in each meal. For instance, pork, chicken and fish for the day 1 and pork, beef, and chicken for the day.

**Figure 1.**
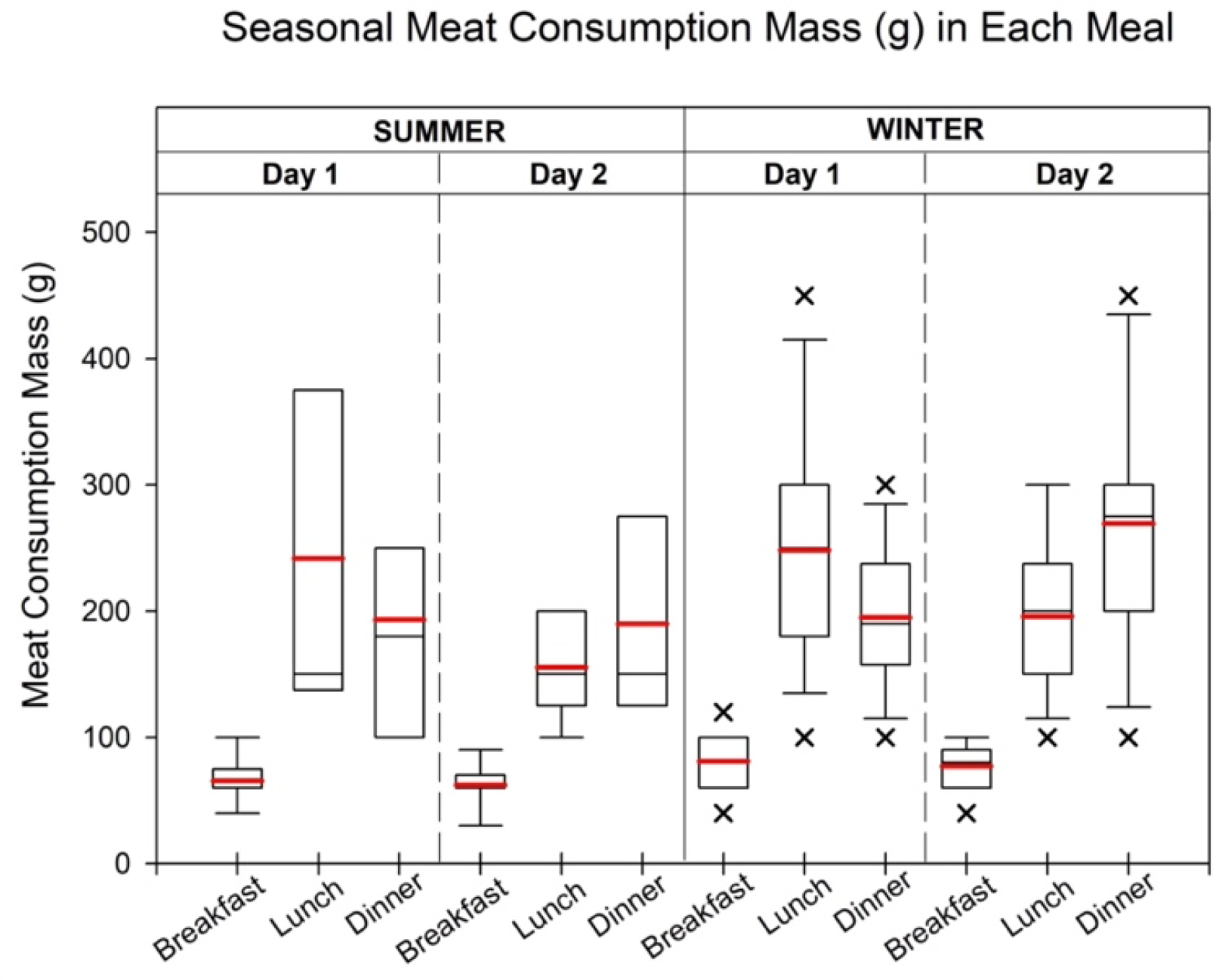
The meat intake volume (g) in each meal at summer and winter. Each of the meat intake time was selected by the highest overall meat consumption in each meal during the summer and winter. Red line represents the mean value of intake. During summer mornings, pork consumption varied between 60g and 90g, averaging around 88g. For lunch on the first day, chicken and beef showed comparable totals, each with a minimum of 100g. Chicken’s maximum consumption, however, was notably higher at 600g. Fish dominated dinner on the first day with a maximum of 400g, while mean fish consumption matched the maximum pork intake. On the second day, chicken consumption (400g) surpassed pork (300g), with similar mean values. Winter consumption mirrored these patterns, with pork exclusive to breakfast (60g minimum), higher average on the first day (81.3g), and chicken and beef prevailing in lunch (minimum 100g).

During the summer season, breakfast consumption of pork showed variations with minimum quantities of 60 g and maximum quantities of 90 g and 80 g, respectively. The average breakfast consumption for each day was approximately 88 g. At lunch on the first day, chicken and beef had comparable total consumption, with minimum values of 100 g for both types. However, chicken had a significantly higher (p<0.05) maximum consumption of 600 g compared to beef. The average consumption of chicken (250 g) was slightly higher than that of beef (200 g). For lunch on the second day, beef consumption at 12 o’clock was lower compared to the consumption at 13 o’clock, although the maximum, mean, and median values (250 g, 190.5 g, and 200 g) were higher during the earlier time period. In terms of dinner on the first day, fish was the most consumed meat type with a maximum quantity of 400g. The mean consumption of fish (200 g) matched the maximum pork consumption. On the second day, chicken consumption (400 g) exceeded pork consumption (300 g), with similar mean and median values for both.

Similar consumption patterns were observed during the winter season, with pork being the only meat consumed during breakfast. The minimum pork consumption remained the same at 60 g for both days, but the average consumption was slightly higher on the first day (81.3 g) compared to the second day (77.1 g). For lunch on each day, chicken and beef were the predominant meat types consumed. On the first day, chicken consumption exceeded fish consumption by 108%, while on the second day, beef consumption was more than four times higher than other meat types. The mean lunch consumption for chicken and beef was 248.1 g and 195.8 g, respectively. During dinner on the first day, fish consumption was more than double that of the second most consumed meat, chicken. However, on the second day, beef consumption was significantly higher. The mean consumption for fish and chicken during dinner was 195 g and 269.2 g, respectively.

Detailed information on dairy product consumption is provided in the Supplementary Materials Table S5. In both summer and winter, the peak consumption of dairy products occurred between 0800 and 0859. The maximum, median, and minimum consumption levels for dairy products during breakfast were 350 g, 60 g, and 50 g or mL, respectively, in summer. In winter, the maximum and minimum consumption remained the same at 300 g or mL, while the median consumption on the second day (140 g or mL) was slightly higher than that of the first day (100 g or mL). Additionally, participants consumed slightly more dairy products on the first day during summer, whereas the trend was reversed in winter.

The meat consumption patterns observed in this study exhibit cultural influences, personal preferences, and seasonal variations. The preference for specific types of meat in each meal align with the findings of the impact of cultural values and beliefs on meat consumption (Hansen et al., 2021). For example, the consistent consumption of pork for breakfast reflects cultural norms, while the higher average consumption of chicken compared to beef may be influenced by perceptions of chicken as a lean and healthy choice (Wang et al., 2023). Additionally, seasonal availability and individual taste preferences contribute to variations in meat consumption. Ueland et al. (2022) found that individuals consume more poultry during winter months when other meat sources may be limited, supporting the higher chicken and beef consumption observed during winter lunches (Ueland, Rødbotten, & Varela, 2022). Spence et al. (2021) also emphasized the role of flavour preferences and seasonal associations, explaining the consistent patterns observed between summer and winter meat consumption (Spence, 2021).

These findings have implications for public health initiatives aiming to promote healthier and sustainable meat consumption. By considering cultural influences, nutritional profiles, and seasonal variations, tailored interventions can be developed. Understanding the complex interplay between individual preferences, cultural norms, and health considerations is crucial. Further research should explore these factors in depth to develop evidence-based strategies. Overall, this study contributes to the growing body of knowledge on meat consumption patterns, informing efforts to promote balanced and sustainable dietary choices.

### 3.3. Water consumption

Water consumption, including water-based drinks such as water, coffee, and tea, exhibited similar patterns throughout both summer and winter periods (Table S6). In summer, the total cumulative daily water intake ranged from 4,398 to 4,899 mL. During the morning hours (07:00 to 11:59), approximately 28.8% and 28.3% of the total water consumption occurred. The period between lunch and dinner (12:00 to 17:59) accounted for approximately 38.5% and 37.6% of the total water intake, while the evening hours (18:00 to 23:59) constituted 32.7% and 34.1% of the total intake. In winter, the total daily water intake ranged from 3,838 to 4,346 mL. Similar to summer, the morning hours accounted for approximately 30.7% and 29.5% of the total water consumption. The period between lunch and dinner represented approximately 40.9% and 40.0% of the total water intake, while the evening hours accounted for 28.4% and 30.5% of the total intake.

Figure 2 presents the maximum, minimum, median, and mean hourly water consumption for each day. In summer, the mean water intake during breakfast, lunch, and dinner on the first day was 241.1 mL, 367.8 mL, and 268.0 mL, respectively. On the second day, the mean water intake slightly increased during dinner compared to the first day, with values of 252.2 mL, 305.7 mL, and 384.4 mL for breakfast, lunch, and dinner, respectively. In winter, the mean volume of water intake during each meal was generally higher than in summer. Specifically, the mean intake during breakfast, lunch, and dinner on the first day was 321.8 mL, 353.6 mL, and 339 mL, respectively. On the following day, the mean intake for breakfast, lunch, and dinner was 315.4 mL, 453.0 mL, and 325.8 mL, respectively. Additional details regarding the maximum, minimum, and median consumption volumes can be found in the Supplementary Materials Table S6.

**Figure 2.**
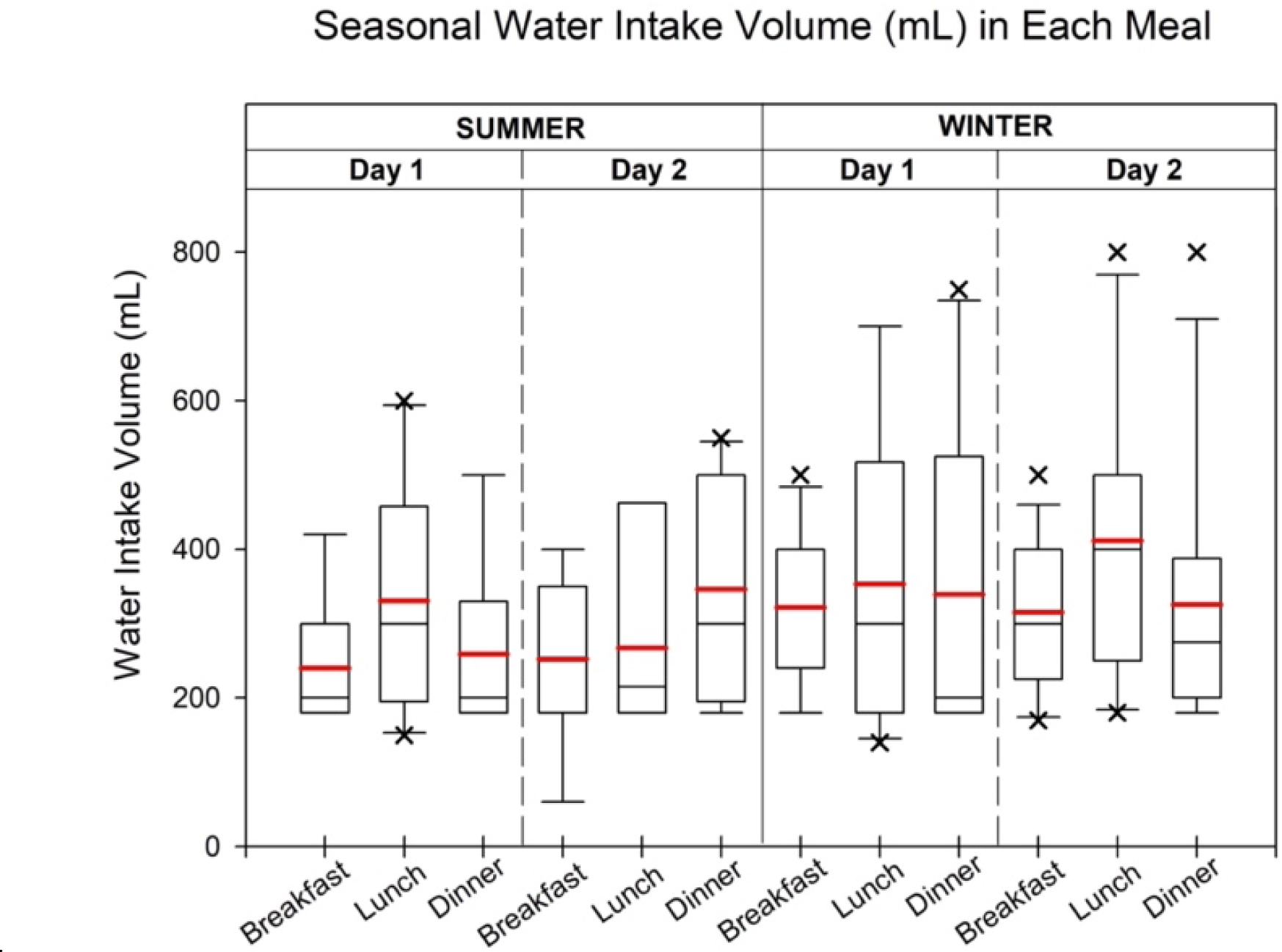
The water intake volume (ml) in each meal at summer and winter. Each of the water intake time was followed by the highest meat consumption in each meal during the summer and winter. Red line represents the mean value of the water intake.

The analysis of water consumption patterns among university faculties revealed consistent trends in both summer and winter, indicating stable hydration practices regardless of the season (Foster et al., 2019; Nishi et al., 2023). These observations align with the findings of previous research on water consumption patterns (Guelinckx et al., 2015; Kenney, Long, Cradock, & Gortmaker, 2015). The consistent water intake during these specific time intervals suggests that individuals prioritize hydration during the morning and lunch hours, possibly to support gut digestion and overall well-being (Vanhaecke, Bretin, Poirel, & Tap, 2022). Adequate water intake during these periods can aid in diluting the concentrations of antibiotics ingested through food, thereby potentially reducing their impact on the gut microbiome (Conlon & Bird, 2014; Zhang, 2022).

In addition to the consistent trends in water consumption, the mean intake volumes during breakfast, lunch, and dinner varied between summer and winter. Higher mean intake volumes in winter may be attributed to the increased thermoregulatory demands during colder months, leading to higher fluid intake (Hosseinlou, Khamnei, & Zamanlu, 2013; Shirreffs, Watson, & Maughan, 2007). This observation is in line with studies that have shown increased water needs in response to environmental factors (Kavouras, 2019). Understanding these variations in water intake throughout the year can inform public health strategies aimed at promoting optimal hydration and managing antibiotic exposure in food-producing animals’ products (Murray et al., 2017).

### 3.4. Antibiotic detection and quantification from the meat samples

Triplicates of 34 food and drink samples were tested to determine the presence of the target antibiotics. All foods mentioned in the survey responses from omnivorous and unknown dietary habit groups in the online survey were included. Most of the detections were over the LOD and LOQ with relatively high accuracy.

Table 2 shows the MRL is the maximum amount of antibiotic residue that is expected to legally remain in food products. ADI is then calculated based on chronic intake of the MRL and a theoretical daily food basket (consisting of 300g meat, 1500mL milk, and 100g eggs). Lastly, TMDI is calculated based on the high quartile bounds of food intake, 65 to 80%, to stress worst-case scenario or conservative limits. Highly consumed antibiotics in table 2 have been detected in all environmental samples including foods and drinking water over the world. The vast majority of those studies reported concentrations which were typically in the ng/L, ng/kg to μg/L or μg/kg range (Fick et al., 2009; Granados-Chinchilla & Rodríguez, 2017; Jammoul & El Darra, 2019; Lucchetti et al., 2004; Patel et al., 2019; Qin et al., 2020; Xu et al., 2021). Although the WHO has limited the concentration of penicillin to be below 100 μg/kg in animal products, greater concentrations were detected in the foods and drinking water (Huang et al., 2020; Jammoul & El Darra, 2019; Okocha, Olatoye, & Adedeji, 2018; Patel et al., 2019; Sachi, Ferdous, Sikder, & Azizul Karim Hussani, 2019). The concentration of amoxicillin and ampicillin in milk were in the range of 28.4 to 96.8 μg/L and in meat were 58.2 to 157 μg/kg (Jammoul & El Darra, 2019). Moreover, the concentration between 0 to 17.8 μg/L of amoxicillin and ampicillin were found in the drinking water (Patel et al., 2019).

**Table 2.**
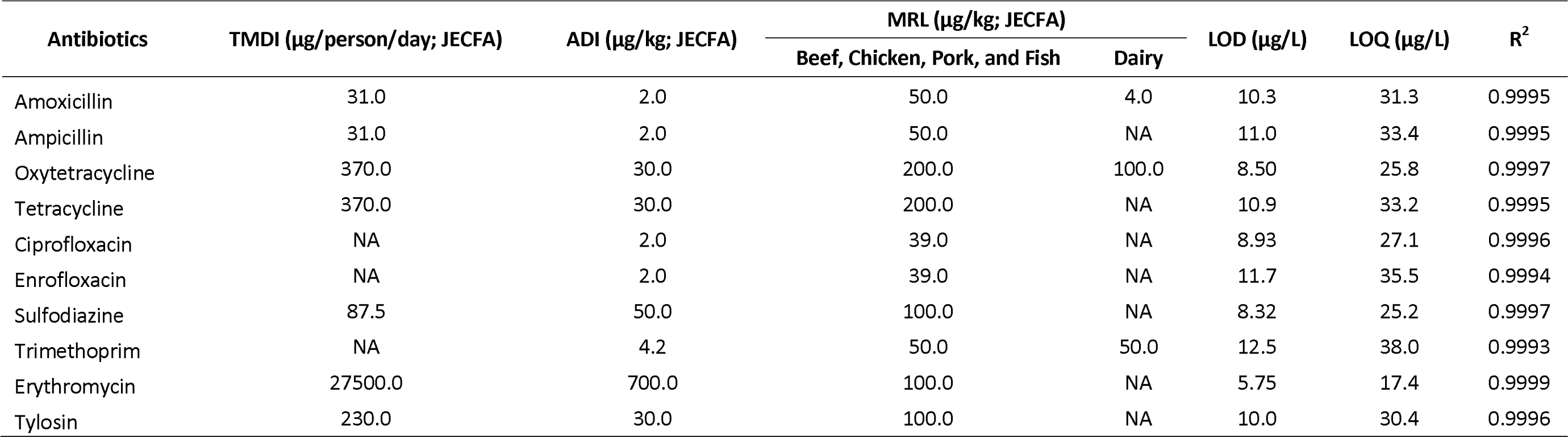
Theoretical maximum daily intake (TMDI), acceptable daily intake (ADI), and Maximum residual level (MRL) of the target antibiotics he Joint FAO/WHO Expert Committee on Food Additives (JECFA), and limit of detection (LOD), limit of quantification (LOQ) and R values of calibration curves for each antibiotic.

Tetracycline and oxytetracycline were detected between 57.0 to 137 μg/L in milk and 82.0 to 691 μg/kg in meat (Granados-Chinchilla & Rodríguez, 2017). In drinking water and tap water, 0.09 to 21.1 μg/L of tetracycline and oxytetracycline were detected (Xu et al., 2021). Long-term consumption of a trace level of tetracyclines needed to be focused on due to its poor biodegradability which may accumulate in the body to make a reservoir of pathogens to have greater resistance. Samples of poultry meats in Europe demonstrated contamination of sulfadiazine and trimethoprim in ranges of 0.64 to 243 μg/kg (Patel et al., 2019). Furthermore, 0.20 to 15.2 μg/L of sulfonamides in drinking water were reported over the world (Qin et al., 2020).

There are no relevant erythromycin and tylosin antibiotic pollution in foods and drinking water data available on these molecules because the usage has been decreased significantly compared to the past decades. However, it has a huge potential to become a high-risk antibiotic as food consumption and production are projected to be increased significantly in South American, Asian, and African countries in the future (Tiseo, Huber, Gilbert, Robinson, & Van Boeckel, 2020). In addition, the FAO designated the concentration of ciprofloxacin and enrofloxacin limit to 2 μg/kg but edible trout still contained 170 to 1006 μg/kg of enrofloxacin in European countries (Lucchetti et al., 2004). Concentration up to 6.5 mg/L of ciprofloxacin was found in drinking water in India (Fick et al., 2009).

In table 3, the concentration of detected antibiotic residues in animal food product samples were calculated based on the survey and chemical analysis results. We have observed 9 of our target antibiotics except erythromycin in samples. Interestingly, processed products such as salami, tuna chunks, ham, meatballs, and sausages exceeded concentration of antibiotics compared to the MRLs. The most exceeded concentration in meat was ENR in sausages (5497.3 μg/kg) which was 141.0 times greater than the MRL (39.0 μg/kg). In addition, the concentration of AMOX in skimmed milk (1481.6 μg/kg) exceeded the MRL by 370.4 times (4 μg/kg). No antibiotics were detected in water samples.

**Table 3.** The detected antibiotics (μg/kg or μg/L) from meat samples including beef, chicken, pork, fish, and dairy products from different ermarket chains in London. Non-detected products (organic salted butter, organic unsalted butter, medium cheddar, dairy spray cream, Greek style yoghurt, sweetened probiotic milk, London tap water, and two different water brand) were not included.

| Type | Name | Concentration of antibiotics (µg/kg or µg/L) |  |  |  |  |  |  |  |  |  |
| --- | --- | --- | --- | --- | --- | --- | --- | --- | --- | --- | --- |
|  |  | TET | OTC | TMP | SDZ | CIP | ENR | AMOX | AMP | TYL | ERY |
| Beef | Ribeye* | - | - | 90.10 | - | - | 616.79 | 674.42 | 1186.64 | - | - |
| Beef | Corned Beef | - | - | 113.74 | - | - | 62.79 | 1940.68 | 271.47 | - | - |
| Beef | Meatballs | - | - | 173.55 | - | - | 2021.48 | - | 348.59 | - | - |
| Beef | Sirloin* | - | - | 88.90 | - | - | 675.16 | 646.48 | 659.73 | - | - |
| Beef | Burger Patty* | - | - | 220.99 | - | - | 1446.31 | 310.89 | 708.35 | - | - |
| Beef | Rump* | - | - | 111.15 | - | - | 451.82 | 775.70 | 988.77 | - | - |
| Beef | Diced Beef* | - | - | 78.57 | - | - | 300.86 | 484.49 | 537.97 | - | - |
| Beef | Minced Beef* | - | - | 264.87 | - | - | 170.3 | 1611.62 | 632.76 | - | - |
| Chicken | Drumsticks* | - | 116.00 | 111.32 | 654.02 | - | - | 1199.00 | - | - | - |
| Chicken | Thighs* | - | - | 197.79 | 1349.36 | - | - | 1535.16 | - | - | - |
| Chicken | Whole Chicken | - | - | 336.22 | 3743.12 | 151.38 | - | - | - | - | - |
| Chicken | Organic Whole Chicken | - | - | 114.12 | 986.95 | 56.78 | - | 1404.57 | - | - | - |
| Chicken | Organic Drumsticks | - | 96.27 | 96.87 | 856.55 | - | - | 1403.14 | - | - | - |
| Chicken | Organic Thighs | - | - | 55.23 | 1028.51 | - | - | 1140.38 | - | - | - |
| Chicken | Chicken Wings | - | - | 67.52 | 674.58 | - | 5976.17 | 589.5 | - | - | - |
| Chicken | Free Range Eggs | - | - | - | - | - | - | 715.58 | - | - | - |
| Chicken | Organic Free-Range Eggs | - | - | - | - | - | - | 818.88 | - | - | - |
| Chicken | Organic Chicken Breast Fillets | - | - | 75.71 | 20.00 | - | - | 233.03 | - | - | - |
| Chicken | Chicken Breast Fillets* | 171.55 | - | 214.79 | 53.19 | 321.31 | - | 1421.10 | - | 327.82 | - |
|  |  | TET | OTC | TMP | SDZ | CIP | ENR | AMOX | AMP | TYL | ERY |
| Dairy | Whole Milk | - | - | 95.92 | - | - | - | - | 36.61 | - | - |
| Dairy | Semi-skimmed Milk* | - | - | 171.34 | - | - | - | 760.04 | - | - | - |
| Dairy | Organic Semi-skimmed Milk | - | - | 96.40 | - | - | - | - | - | - | - |
| Dairy | Skimmed Milk | - | - | 288.82 | - | - | - | 481.62 | - | - | - |
| Fish | Mackerel Fillets | 391.00 | 374.50 | - | - | - | - | - | - | - | - |
| Fish | Salmon Fillets | - | - | 191.21 | - | - | - | - | 415.78 | - | - |
| Fish | Tuna Chunks in Sunflower Oil | - | - | 76.39 | 765.34 | - | - | 2968.48 | - | - | - |
| Fish | Cod Fillets | 648.25 | 1298.77 | - | - | - | 205.42 | - | - | - | - |
| Fish | Haddock Fillets | 279.27 | 220.61 | - | - | - | 538.61 | - | - | - | - |
| Pork | Salami Slices* | - | - | 81.25 | - | 425.09 | 4220.61 | 6866.87 | - | - | - |
| Pork | Pork Sausages* | - | - | - | - | - | 5497.34 | 216.67 | - | - | - |
| Pork | Salty Canned Pork | - | - | 184.56 | 77.91 | - | - | - | - | - | - |
| Pork | Smoked Streaky Bacon Rashers | - | - | 222.18 | - | - | - | - | - | - | - |
| Pork | Unsmoked Streaky Bacon Rashers | - | - | 120.02 | - | - | - | - | - | - | - |
| Pork | British Pork Ribs | - | - | 170.53 | 34.80 | - | - | - | - | - | - |
| Pork | British Pork Chops | - | - | 461.68 | 116.19 | - | - | 1616.09 | - | - | - |
| Pork | British Pork Belly Slices | - | - | 107.55 | - | - | - | - | - | - | - |
| Pork | British Pork Loin | - | - | 185.42 | 1117.7 | - | - | - | - | - | - |
| Pork | Smoked Back Bacon Rashers | - | - | 123.07 | - | - | 61.68 | - | - | - | - |
| Pork | Unsmoked Back Bacon Rashers | - | - | 156.97 | - | - | 37.24 | - | - | - | - |
| Pork | Ham Slices* | - | 71.83 | 305.11 | - | - | - | 2387.85 | - | - | - |

\*Products that exceed ADI level of amoxicillin, ampicillin and enrofloxacin and selected for the EMI calculation based on the diet survey.

#### 3.4.1. Beef and dairy products

In beef, 8 different products were analysed including ribeye, sirloin, rump, diced beef, minced beef, corned beef, burger patty, and meatballs. AMOX, AMP, ENR, and TMP were commonly detected from all beef products, while AMOX was not detected in meatballs. All the detected concentrations were greater than MRLs. Ten different dairy products were tested but there were no target antibiotics detected in 6 of the products including organic salted butter, organic unsalted butter, medium cheddar, dairy spray cream, Greek-style yoghurt, and sweetened yoghurt drink. However, β-lactams and TMP were commonly found in the remaining 4 products (Whole milk, semi-skimmed milk, organic semi-skimmed milk, and skimmed milk). AMP in whole milk was detected below the MRL, but concentrations of AMOX 29.6 times higher than the MRL were detected in skimmed milk.

Cattle are routinely given antibiotics to treat and prevent mastitis which is an infection of the udder and very common (Smith et al., 1998). It can be subclinical, where they show no obvious symptoms, or clinal which causes painful swellings in one or more quarters of the udder. Mastitis is commonly treated with antibiotics administered as an intramammary directly into the cow’s teat (Holman, Yang, & Alexander, 2019). There are various types of mastitis-causing organisms including staphylococci. Antibiotics used to treat *Staphylococcus aureus* mastitis including, AMOX, AMP, ERY, TYL, ENR, and TMP, which it makes highly probable with our antibiotic detection from beef and dairy products (Rayner & Munckhof, 2005) (Table 3). Furthermore, mastitis infection can be monitored in herds through cell counts in milk and farmers are financially penalised by dairy companies for high cell counts (EU, 2017). Milk from infected cows must be withheld from sale for the required withdrawal periods. However, the fermented or intensively processed products such as yoghurt, cheese, cream, and butter were prevented to use antibiotics and put extra-care with the antibiotic detections for the higher rate of fermentation or cost-effectiveness (Erdogan, Gurses, Turkoglu, & Sert, 2001). It explains why the milk had antibiotic residues while no detections from fermenting-based products.

#### 3.4.2. Chicken

In chicken, 11 different products consisting of 5 organic products and 6 conventional products were tested. In general, AMOX, SDZ, and TMP were detected from the products. All OTC and TET detections were lower than MRLs. CIP concentrations from the chicken breast were also lower than the MRL. ENR in BBQ chicken wings (5492.3 μg/kg) was the highest concentration detected from all samples and it exceeded MRL by 153.2 times. Interestingly, 4 out of 5 organic products (drumstick, chicken breast, eggs, and whole chicken) had higher concentrations of antibiotics compared to the same types of conventional products. The organic drumstick, chicken breast, and egg had 17.0, 509.9, and 14.4% higher concentration of AMOX compared to the conventional products, respectively. Also, 31.0% higher concentration of SDZ was detected in organic drumstick. 1404.6 μg/kg of AMOX was detected at organic whole chicken while AMOX was not detected in the conventional product.

The poultry industry is split into two parts: the broiler industry, which produces birds slaughtered at 6 to 7 weeks old for the table, and the egg-producing sector where layers are reared and placed in battery cages at 16 to 18 weeks for one egg laying cycle and then killed. Treatment is required for any outbreak of necrotic enteritis, *Colispticaemia salmonellosis* causing mortality, outbreak of mycoplasma infection or outbreak of necrotic dermatitis (*Staphylococcus aureus*) (Christian, Vivian Etsiapa, Crystal Ngofi, & Frank Boateng, 2018). The antibiotics used for salmonella and *E. coli* may include ENR and AMOX, which are reflecting to our results in table 3. We can assume that CIP is metabolized from ENR, and it is important to regulate the intensive usage of ENR because of the incidence of isolates of multi drug-resistant *Salmonella typhimurium DT104* from humans which are resistant to CIP (Weill et al., 2006).

#### 3.4.3. Pork

We had the highest variety of food products from pork, 12, and all the products contained TMP except sausages. Although TMP was the most frequently detected, lipophilic antibiotics such as AMOX and ENR were detected at concentrations exceeding MRL. For instance, 137.3 and 108.2 times higher AMOX and ENR than its MRLs were measured in salami, and 141.0 times higher concentration of ENR was detected in sausage. The concentration of OTC in ham, ENR in unsmoked back bacon, SDZ in ribs and Salty Canned Pork were lower than respective MRLs.

Swine is reared indoors receive intensive antibiotic treatment throughout their life until slaughter, usually at under 6 months old. Most conventional herds are watered or fed with the growth promoters during the early stages of growth. It is true that we did not detect any of growth promotors from pork products (Table 3), but there are various growth promoter antibiotics used in pig farming that are more cost-effective compared to our macrolides. For instance, avilamycin, carbadox, flavomycin, olaquindox, spiramycin, and salinomycin (Lekagul, Tangcharoensathien, & Yeung, 2019). Our detection could be explained with the most conventional herds which their antibiotic treatment starts soon after birth. Piglets are typically treated with AMP, ENR, TMP and SDZ for *E. coli* enteritis and for respiratory disease (Rhouma, Fairbrother, Beaudry, & Letellier, 2017) and slaughtered after 6 months.

#### 3.4.4. Fish

Five different farmed and wild fish products were tested and OTC, TET and ENR were commonly detected from wild fish including mackerel, cod, and haddocks, while β-lactam and sulfonamides were measured in farmed fish such as salmon and tuna. All the measured concentrations were above the MRLs. ENR was measured at 13.8 times higher than the MRL from haddock fillets, and AMOX was detected at a concentration 59.4 times higher compared to its MRL.

Fish farming products are still contaminated with antibiotics with relatively high concentrations, though the official usage of antibiotics has been significantly reduced from the past due to increased regulation, vaccination, and the segregation of farmed fish by age (Ma, Bruce, Jones, & Cain, 2019). Recent studies have raised concerns that antibiotics enter fish farms not only direct medication, but also feeding with chicken faeces which treated with intensive treatments (Elsaidy, Abouelenien, & Kirrella, 2015; Ke et al., 2020). In table 3, it is not unreasonable to postulate that AMOX, SDZ, and TMP concentrations from chicken were similar to the farmed fishes such as salmon and tuna. Moreover, a large number of feed pellets were found in the gut contents of wild fishes such as mackerel, cod, and haddocks near a fish farm in Scotland (Black, Hansen, & Holmer, 2008). Furthermore, wild fishes are more vulnerable to antibiotic aquatic pollution which is rarely taken into consideration (Naylor et al., 2021). It is worth noting that most of the antibiotics used are persistent in the environment and spread from the farms to surrounding areas where accumulation in sediments may occur (Devarajan et al., 2015). Residues of antibiotic concentrations may far exceed levels accepted for human consumption (Black et al., 2008). In addition, fishes were also treated with antibiotics after being caught from the ocean to avoid the pathogenic penalties of regulation (Booth, Aga, & Wester, 2020).

### 3.5. The elimination of antibiotics during cooking processes

The application of veterinary antibiotics to food-producing animals has led to residues occurring in the food products such as beef, chicken, pork, and dairy products which increases the risks to human health. In addition, the removal of antibiotics in drinking water is highly variable depending on treatment technologies, including activated carbon adsorption, ozonation, membrane filtration, and advanced oxidation process (AOPs) (Reungoat, Escher, Macova, & Keller, 2011). According to Liu et al. (2015), removal of antibiotics was effective using a combination of activated carbon adsorption and ozonation in water treatment process (Liu et al., 2016). Sand biofiltration is expected to be widely demanding technology because of the low cost of sand (Paredes, Fernandez-Fontaina, Lema, Omil, & Carballa, 2016). AOPs such as Fenton oxidation and photocatalytic oxidation, demonstrated high efficiencies of antibiotic removal (>90%), however, formation of various by-products of antibiotic are the main concern of AOPs (Luo et al., 2014). Properties of antibiotics including pharmacokinetic characteristics, physicochemical or biological processes, and improper usages are considered as factors influencing the occurrence of antibiotic residues in foods and drinking water (Manyi-Loh et al., 2018).

Most hygiene guidance states that foods should be kept above boiling point for enough time to kill the harmful pathogens (US Government, 2023). Although the majority of pathogens are killed in the cooking process, studies have found that the concentration of antibiotic residues in foods was not significantly degraded after cooking at a temperature above 100°C for more than 30 mins (Shaltout, 2019). Different cooking practices, including boiling, frying, and grilling, at different periods of cooking time were examined to understand antibiotic concentration reduction in foods (Abou-Raya, Shalaby, Salama, & Mehaya, 2013; Moats, 1999; Shalaby, Salama, Abou-Raya, Emam, & Mehaya, 2011). Firstly, tetracyclines, including oxytetracycline, tetracycline, chlortetracycline, and doxycycline, were tested to determine the reduction of antibiotic concentrations with different cooking procedures (boiling, microwave, and roasting) at different time ranges (0, 10, 15, 20, 30, 40, 60, and 80 minutes (Abou-Raya et al., 2013). A significant reduction of chlortetracycline and doxycycline concentration was observed with all cooking procedures from 30 mins above. However, oxytetracycline and tetracycline were not reduced by more than 50% at the maximum exposure time, 80 mins. In addition, the concentration of chlortetracycline and oxytetracycline reduced by 27.6% and 35.6% after boiling milk for 30 mins, respectively (Moats, 1999). Also, 11.1% of antibiotics were inactivated by heating for 30 mins in water(Moats, 1999).

Furthermore, degrading antibiotics, including ciprofloxacin, tylosin, oxytetracycline, sulfonamides, in beef, chicken, and rabbit meat samples were ineffective to reduce the concentration in the meat samples by boiling and roasting processes (Fahim, Shaltout, & El shatter, 2019; Salama, Abou-Raya, Shalaby, Emam, & Mehaya, 2011). For instance, ciprofloxacin was reduced by 17.0% and 22.4% after roasting and boiling chicken muscle for 30 mins, respectively (Fahim et al., 2019). Furusawa and Hanabusa (2002) have tested the degradation effect of boiling, roasting and microwaving to sulfonamides in chicken muscle (Furusawa & Hanabusa, 2002). Sulfadiazine was appeared to be stable in boiling, roasting and microwaving methods with showing 32.3% reduction at maximum compared to other sulfonamides, including sulfamethoxazole, sulfamonomethoxine, and sulfaquinoxaline (45.0-61.0%). In addition, degradation of tylosin in chicken meatball under microwaving had a least reduction (2.8%) compared to the other antibiotics while the microwaving was showing a strong antibiotic reduction (Salaramoli, Heshmati, Kamkar, & Hassan, 2015). Interestingly, the concentration of enrofloxacin in raw meat samples were increased by 44.0-310% during grilling and roasting due to the loss of moisture content in the samples (Sobral, Cunha, Faria, & Ferreira, 2018).

### 3.6. Estimated Meal Intake (EMI) of antibiotics from each meal

The estimation of antibiotic residues consumed via each meal provides valuable insights into the potential exposure of individuals to these antimicrobial agents. In this study, the estimated daily intake formula modified from the US FDA was utilized to calculate the antibiotic intake from each meal. The formula took into account factors such as portion size, concentration of antibiotics in the consumed foods, and the frequency of consumption. Moreover, the total volume of the gut juice in stomach and duodenum was set to 60 mL based on the monitoring human digestive system using a magnetic resonance imaging quantification (Murray et al., 2017). By applying these parameters, the study aimed to estimate the antibiotics that individuals may be exposed to during specific meals.

The results showed varying levels of estimated antibiotic residues in different meals. In table 4, during the first day, the estimated antibiotic intake from breakfast included 141.3 mg/L of amoxicillin and 64.2 mg/L of enrofloxacin, while lunch had an estimated intake of 399.4 mg/L of amoxicillin. Notably, dinner on the first day was estimated to be below the acceptable daily intake (ADI) concentration for all antibiotics analysed. Similarly, the estimated antibiotic residues from each meal on the second day showed variations, with breakfast, lunch, and dinner containing different antibiotic concentrations.

**Table 4.** The average of estimated antibiotic intake per meal (mg/L) during the summer and winter using EMI equation based on the survey results.

| Day | Meals | Estimated Antibiotics (mg/L) |  |  |
| --- | --- | --- | --- | --- |
|  |  | Amoxicillin | Ampicillin | Enrofloxacin |
| 1st | Breakfast | 141.3 | - | 64.2 |
|  | Lunch | 399.4 | - | - |
|  | Dinner | <ADI<sup>a</sup> | <ADI | - |
| 2nd | Breakfast | 132.5 | <ADI | 80.3 |
|  | Lunch | 170.7 | 193.5 | 194.4 |
|  | Dinner | 408.1 | - | - |

<sup>a</sup> Acceptable Daily Intake

It is essential to consider the potential health implications of consuming estimated antibiotic residues through meals. Antibiotic residues in food can contribute to the development and spread of antibiotic resistance, posing a significant public health concern. The ingestion of antibiotic residues can impact the gut microbiota, potentially disrupting its balance and affecting overall gut health. Furthermore, exposure to subtherapeutic levels of antibiotics through food consumption may contribute to the selection and proliferation of antibiotic-resistant bacteria in the gut.

Recent research has highlighted the importance of monitoring and minimizing antibiotic residues in food. A study by Arsène et al. (2022) investigated the presence of antibiotic residues in animal-derived food products and assessed their potential impact on human health (Arsène et al., 2022). The findings underscored the need for strict monitoring and regulation of antibiotic residues in food to mitigate the risks associated with antibiotic resistance development.

The estimation of antibiotic residues consumed via each meal provides valuable information on the potential exposure of individuals to these antimicrobial agents. The results of this study demonstrate variations in estimated antibiotic residues in different meals, emphasizing the importance of monitoring and minimizing antibiotic residues in food to mitigate the risks associated with antibiotic resistance. Further research is warranted to evaluate the long-term effects of consuming estimated antibiotic residues and develop strategies to ensure food safety and preserve the effectiveness of antibiotics in the treatment of bacterial infections.

Investigating the luminal concentration of prescribed pharmaceuticals, particularly antibiotics, within the human digestive system presents unique challenges due to the complex dynamics of gastrointestinal environments. Previous research attempted to estimate luminal prescribed pharmaceutical concentrations by considering gut environmental conditions and geomatic assumptions (Korsten, Smits, Garssen, & Vromans, 2019; Thursby & Juge, 2017). Notably, the uneven distribution of prescribed antibiotics within chyme or bolus formations in the small intestine leads to variations in antibiotic concentrations in different pockets (Korsten et al., 2019). Even when assuming an even distribution throughout the bolus, luminal antibiotic concentrations calculated based on minimum prescription dosages still surpass the estimated concentrations presented in Table 4. For instance, the minimum prescription dose of amoxicillin in the UK’s NHS is 250 mg, which, when divided by the volume of a meal, yields concentrations around four times greater than our estimated maximum luminal antibiotic concentration. Similar calculations for ampicillin and enrofloxacin also demonstrate notable discrepancies between prescribed dosages and estimated luminal concentrations.

*Lactobacillus sp., Escherichia coli and Enterococcus spp.* are the most predominant species in human duodenum (Angelakis et al., 2015). Considering the human gut microbiota’s susceptibility to antibiotics, it is noteworthy that different bacterial species exhibit varying MICs. According to the EUCAST MIC dataset, *Lactobacillus sp*. displayed MIC levels of up to 32 mg/L for ampicillin. *E. coli* exhibited the highest MIC levels for amoxicillin and ampicillin, reaching up to 512 mg/L, and 8 mg/L for enrofloxacin. Meanwhile, *Enterococcus spp.* reported MIC levels of up to 256 mg/L for amoxicillin and ampicillin. Although enrofloxacin MIC data for *Enterococcus spp.* is unavailable, ciprofloxacin, the main metabolite of enrofloxacin, showed a maximum MIC level of 512 mg/L. In addition to MIC levels, the duration of antibiotic exposure is also critical. For example, to eradicate *E. coli*, one to three hours of antibiotic exposure is typically required, whereas the bacteria reproduce approximately every 20 mins (Korsten et al., 2019). With a duodenal transition time of around 18 mins, which is a third of the minimum time required to eliminate *E. coli*, it is likely that these bacteria would survive chronic exposure to sub-therapeutic antibiotic levels.

While the estimated luminal antibiotic exposure from dietary sources in our study may not reach levels sufficient to eradicate gut microbiota, understanding the potential consequences of chronic luminal exposure is crucial to assessing risks associated with antimicrobial resistance development. Future investigations should delve into the intricate interplay between chronic luminal antibiotic exposure and gut microbial communities to provide comprehensive insights into the possible implications on antimicrobial resistance development and overall gut health.

## Conclusions

In conclusion, this study underscores the potential risks of dietary antibiotic exposure, even at sub-therapeutic levels, in contributing to the development of antibiotic resistance. The comparison of estimated luminal antibiotic concentrations from meals with prescribed dosages highlights substantial differences, raising concerns about the efficacy of dietary antibiotics. Additionally, the analysis of minimum inhibition concentrations for key gut bacteria emphasizes the complexity of microbial responses. Although dietary exposure may not achieve eradication levels, the chronic exposure to sub-therapeutic concentrations could foster antimicrobial resistance. This underscores the urgency for stringent regulation of antibiotic residues in food and a deeper investigation into the long-term impacts of chronic luminal antibiotic exposure on gut microbiota and antimicrobial resistance development.

## Data Availability

All data produced in the present study are available upon reasonable request to the authors.

## Acknowledgements

We thank to the survey respondents who voluntarily participated in this study for providing valuable data during the pandemic.

## Declaration of interests

There are no known competing financial interests or personal relationships that could have appeared to influence the work reported in this paper.

## Author Contributions

**Jegak Seo:** Conceptualization, Methodology, Validation, Formal analysis, Investigation, Writing-Original Draft, Writing - Review & Editing, Visualization, Project administration, Funding acquisition; **Frank Kloprogge**: Conceptualization, Methodology, Writing - Review & Editing, Supervision, Project administration; **Andrew M. Smith**: Methodology, Validation, Writing - Review & Editing, Supervision; **Kersti Karu:** Methodology, Validation, Formal analysis, Investigation, Resources, Writing - Review & Editing; **Lena Ciric**: Conceptualization, Methodology, Validation, Formal analysis, Investigation, Resources, Writing - Review & Editing, Supervision, Project administration, Funding acquisition

## Notes

### Competing Interest Statement

The authors have declared no competing interest.

### Funding Statement

This study did not receive any funding.

### Author Declarations

Ethics Committee of the University College London gave ethical approval for this work, project number [19139/001].

## Reference

Abou-Raya, S., Shalaby, A. R., Salama, N., & Mehaya, F. (2013). Effect of Ordinary Cooking Procedures on Tetracycline Residues in Chicken Meat. Journal of Food and Drug Analysis, 21, 80–86.

Ali Mirza, S., Afzaal, M., Begum, S., Arooj, T., Almas, M., Ahmed, S., & Younus, M. (2020). Chapter 11 - Uptake mechanism of antibiotics in plants. In M. Z. Hashmi (Ed.), Antibiotics and Antimicrobial Resistance Genes in the Environment (Vol. 1, pp. 183–188): Elsevier.

Andrew Bamidele, F., & Oluwakamisi Festus, A. (2019). Veterinary Drug Residues in Meat and Meat Products: Occurrence, Detection and Implications. In B. Samuel Oppong, S. Mani, A. Reimmel Kwame & P. K. Ramkumar (Eds.), Veterinary Medicine and Pharmaceuticals (pp. Ch. 5). Rijeka: IntechOpen.

Angelakis, E., Armougom, F., Carrière, F., Bachar, D., Laugier, R., Lagier, J.-C., Robert, C., Michelle, C., Henrissat, B., & Raoult, D. (2015). A Metagenomic Investigation of the Duodenal Microbiota Reveals Links with Obesity. PLOS ONE, 10 (9), e0137784.

Arsène, M. M. J., Davares, A. K. L., Viktorovna, P. I., Andreevna, S. L., Sarra, S., Khelifi, I., & Sergueïevna, D. M. (2022). The public health issue of antibiotic residues in food and feed: Causes, consequences, and potential solutions. Vet World, 15 (3), 662–671.

Ayukekbong, J. A., Ntemgwa, M., & Atabe, A. N. (2017). The threat of antimicrobial resistance in developing countries: causes and control strategies. Antimicrobial Resistance & Infection Control, 6 (1), 47.

Black, K. D., Hansen, P. K., & Holmer, M. (2008). Salmon Aquaculture Dialogue: Working Group Report on Benthic Impacts and Farm Siting. In (pp. 54): Scottish Association for Marine Science, Oban, Scotland, Institute for Marine Science, Bergen, Norway, University of Southern Denmark, Odense, Denmark.

Booth, A., Aga, D. S., & Wester, A. L. (2020). Retrospective analysis of the global antibiotic residues that exceed the predicted no effect concentration for antimicrobial resistance in various environmental matrices. Environment International, 141, 105796.

Christian, A., Vivian Etsiapa, B., Crystal Ngofi, Z., & Frank Boateng, O. (2018). Antibiotic Use in Poultry Production and Its Effects on Bacterial Resistance. In K. Yashwant (Ed.), Antimicrobial Resistance (pp. Ch. 3). Rijeka: IntechOpen.

Conlon, M. A., & Bird, A. R. (2014). The impact of diet and lifestyle on gut microbiota and human health. Nutrients, 7 (1), 17–44.

Devarajan, N., Laffite, A., Graham, N. D., Meijer, M., Prabakar, K., Mubedi, J. I., Elongo, V., Mpiana, P. T., Ibelings, B. W., Wildi, W., & Poté, J. (2015). Accumulation of clinically relevant antibiotic-resistance genes, bacterial load, and metals in freshwater lake sediments in Central Europe. Environ Sci Technol, 49 (11), 6528–6537.

Economou, V., & Gousia, P. (2015). Agriculture and food animals as a source of antimicrobial-resistant bacteria. Infect Drug Resist, 8, 49–61.

Elsaidy, N., Abouelenien, F., & Kirrella, G. A. K. (2015). Impact of using raw or fermented manure as fish feed on microbial quality of water and fish. The Egyptian Journal of Aquatic Research, 41 (1), 93–100.

Erdogan, A., Gurses, M., Turkoglu, H., & Sert, S. (2001). Fixing the time of the milk ripening depending on the content of immobilized johourt ferment. Pakistan Journal of Biological Sciences, 4 (7), 886–887.

EU. (2017). Health and Food Audits and Analysis Programme 2017. In (pp. 57). Luxemburg: European Commission.

Fahim, H., Shaltout, F., & El shatter, M. A. (2019). Evaluate antibiotic residues in beef and effect of cooking and freezing on it. Benha Veterinary Medical Journal.

FAO. (2016). Drivers, dynamics and epidemiology of antimicrobial resistance in animal production. Rome.

FAO. (2018). Dietary Assessment: A resource guide to method selection and application in low resource settings. Rome: Food and Agriculture Organization of the United Nations.

Fda, U. (2018). Guidance for Industry: Estimating Dietary Intake of Substances in Food. In U. Fda (Ed.).

Fick, J., Söderström, H., Lindberg, R. H., Phan, C., Tysklind, M., & Larsson, D. G. (2009). Contamination of surface, ground, and drinking water from pharmaceutical production. Environ Toxicol Chem, 28 (12), 2522–2527.

Foster, E., Lee, C., Imamura, F., Hollidge, S. E., Westgate, K. L., Venables, M. C., Poliakov, I., Rowland, M. K., Osadchiy, T., Bradley, J. C., Simpson, E. L., Adamson, A. J., Olivier, P., Wareham, N., Forouhi, N. G., & Brage, S. (2019). Validity and reliability of an online self-report 24-h dietary recall method (Intake24): a doubly labelled water study and repeated-measures analysis. J Nutr Sci, 8, e29.

Furusawa, N., & Hanabusa, R. (2002). Cooking effects on sulfonamide residues in chicken thigh muscle. Food Research International, 35, 37–42.

Gaal, S., Kerr, M. A., Ward, M., McNulty, H., & Livingstone, M. B. E. (2018). Breakfast Consumption in the UK: Patterns, Nutrient Intake and Diet Quality. A Study from the International Breakfast Research Initiative Group. Nutrients, 10 (8).

Government, U. (2021). National Diet and Nutrition Survey: Diet, nutrition and physical activity in 2020: A follow up study during COVID-19. In P. H. England & F. S. Agency (Eds.), (pp. 54).

Government, U. (2023). Cook to a Safe Minimum Internal Temperature. In F. Safety (Ed.). Granados-Chinchilla, F., & Rodríguez, C. (2017). Tetracyclines in Food and Feedingstuffs: From Regulation to Analytical Methods, Bacterial Resistance, and Environmental and Health Implications. J Anal Methods Chem, 2017, 1315497.

Guelinckx, I., Iglesia, I., Bottin, J. H., De Miguel-Etayo, P., González-Gil, E. M., Salas-Salvadó, J., Kavouras, S. A., Gandy, J., Martinez, H., Bardosono, S., Abdollahi, M., Nasseri, E., Jarosz, A., Ma, G., Carmuega, E., Thiébaut, I., & Moreno, L. A. (2015). Intake of water and beverages of children and adolescents in 13 countries. Eur J Nutr, 54 Suppl 2 (Suppl 2), 69–79.

Hansen, J., Sparleanu, C., Liang, Y., Büchi, J., Bansal, S., Caro, M. Á., & Staedtler, F. (2021). Exploring cultural concepts of meat and future predictions on the timeline of cultured meat. Future Foods, 4, 100041.

Holman, D. B., Yang, W., & Alexander, T. W. (2019). Antibiotic treatment in feedlot cattle: a longitudinal study of the effect of oxytetracycline and tulathromycin on the fecal and nasopharyngeal microbiota. Microbiome, 7 (1), 86.

Hosseinlou, A., Khamnei, S., & Zamanlu, M. (2013). The effect of water temperature and voluntary drinking on the post rehydration sweating. Int J Clin Exp Med, 6 (8), 683–687.

Huang, L., Mo, Y., Wu, Z., Rad, S., Song, X., Zeng, H., Bashir, S., Kang, B., & Chen, Z. (2020). Occurrence, distribution, and health risk assessment of quinolone antibiotics in water, sediment, and fish species of Qingshitan reservoir, South China. Scientific Reports, 10 (1), 15777.

Jammoul, A., & El Darra, N. (2019). Evaluation of Antibiotics Residues in Chicken Meat Samples in Lebanon. Antibiotics (Basel), 8 (2).

Kavouras, S. A. (2019). Hydration, dehydration, underhydration, optimal hydration: are we barking up the wrong tree? Eur J Nutr, 58 (2), 471–473.

Ke, F., Gao, Y., Liu, L., Zhang, C., Wang, Q., & Gao, X. (2020). Comparative analysis of the gut microbiota of grass carp fed with chicken faeces. Environ Sci Pollut Res Int, 27 (26), 32888–32898.

Kenney, E. L., Long, M. W., Cradock, A. L., & Gortmaker, S. L. (2015). Prevalence of Inadequate Hydration Among US Children and Disparities by Gender and Race/Ethnicity: National Health and Nutrition Examination Survey, 2009-2012. Am J Public Health, 105 (8), e113–118.

Klein, E. Y., Van Boeckel, T. P., Martinez, E. M., Pant, S., Gandra, S., Levin, S. A., Goossens, H., & Laxminarayan, R. (2018). Global increase and geographic convergence in antibiotic consumption between 2000 and 2015. Proceedings of the National Academy of Sciences, 115 (15), E3463–E3470.

Korsten, S. G. P. J., Smits, E. A. W., Garssen, J., & Vromans, H. (2019). Modeling of the luminal butyrate concentration to design an oral formulation capable of achieving a pharmaceutical response. PharmaNutrition, 10, 100166.

Kraemer, S. A., Ramachandran, A., & Perron, G. G. (2019). Antibiotic Pollution in the Environment: From Microbial Ecology to Public Policy. Microorganisms, 7 (6).

Lekagul, A., Tangcharoensathien, V., & Yeung, S. (2019). Patterns of antibiotic use in global pig production: A systematic review. Veterinary and Animal Science, 7, 100058.

Liu, J., Sun, Q., Zhang, C., Li, H. a., Song, W., Zhang, N. i., & Jia, X. (2016). Removal of typical antibiotics in the advanced treatment process of productive drinking water. Desalination and Water Treatment, 57 (24), 11386–11391.

Llor, C., & Bjerrum, L. (2014). Antimicrobial resistance: risk associated with antibiotic overuse and initiatives to reduce the problem. Ther Adv Drug Saf, 5 (6), 229–241.

Lucchetti, D., Fabrizi, L., Guandalini, E., Podestà, E., Marvasi, L., Zaghini, A., & Coni, E. (2004). Long depletion time of enrofloxacin in rainbow trout (Oncorhynchus mykiss). Antimicrob Agents Chemother, 48 (10), 3912–3917.

Luo, Y., Guo, W., Ngo, H. H., Nghiem, L. D., Hai, F. I., Zhang, J., Liang, S., & Wang, X. C. (2014). A review on the occurrence of micropollutants in the aquatic environment and their fate and removal during wastewater treatment. Science of The Total Environment, 473-474, 619–641.

Ma, J., Bruce, T. J., Jones, E. M., & Cain, K. D. (2019). A Review of Fish Vaccine Development Strategies: Conventional Methods and Modern Biotechnological Approaches. Microorganisms, 7 (11).

Manyi-Loh, C., Mamphweli, S., Meyer, E., & Okoh, A. (2018). Antibiotic Use in Agriculture and Its Consequential Resistance in Environmental Sources: Potential Public Health Implications. Molecules, 23 (4).

Moats, W. A. (1999). The effect of processing on veterinary residues in foods. Adv Exp Med Biol, 459, 233–241.

Morehead, M. S., & Scarbrough, C. (2018). Emergence of Global Antibiotic Resistance. Prim Care, 45 (3), 467–484.

Murray, K., Hoad, C. L., Mudie, D. M., Wright, J., Heissam, K., Abrehart, N., Pritchard, S. E., Al Atwah, S., Gowland, P. A., Garnett, M. C., Amidon, G. E., Spiller, R. C., Amidon, G. L., & Marciani, L. (2017). Magnetic Resonance Imaging Quantification of Fasted State Colonic Liquid Pockets in Healthy Humans. Molecular Pharmaceutics, 14 (8), 2629–2638.

Naylor, R. L., Hardy, R. W., Buschmann, A. H., Bush, S. R., Cao, L., Klinger, D. H., Little, D. C., Lubchenco, J., Shumway, S. E., & Troell, M. (2021). A 20-year retrospective review of global aquaculture. Nature, 591 (7851), 551–563.

Nishi, S. K., Babio, N., Paz-Graniel, I., Serra-Majem, L., Vioque, J., Fitó, M., Corella, D., Pintó, X., Bueno-Cavanillas, A., Tur, J. A., Diez-Ricote, L., Martinez, J. A., Gómez-Martínez, C., González-Botella, A., Castañer, O., Alvarez-Sala, A., Montesdeoca-Mendoza, C., Fanlo-Maresma, M., Cano-Ibáñez, N., Bouzas, C., Daimiel, L., Zulet, M. Á., Sievenpiper, J. L., Rodriguez, K. L., Vázquez-Ruiz, Z., & Salas-Salvadó, J. (2023). Water intake, hydration status and 2-year changes in cognitive performance: a prospective cohort study. BMC Medicine, 21 (1), 82.

O’Neill, J. (2016). Tackling drug-resistant infections globally: final report and recommendations: Government of the United Kingdom.

Okocha, R. C., Olatoye, I. O., & Adedeji, O. B. (2018). Food safety impacts of antimicrobial use and their residues in aquaculture. Public Health Reviews, 39 (1), 21.

Paredes, L., Fernandez-Fontaina, E., Lema, J. M., Omil, F., & Carballa, M. (2016). Understanding the fate of organic micropollutants in sand and granular activated carbon biofiltration systems. Sci Total Environ, 551-552, 640–648.

Patel, M., Kumar, R., Kishor, K., Mlsna, T., Pittman, C. U., Jr., & Mohan, D. (2019). Pharmaceuticals of Emerging Concern in Aquatic Systems: Chemistry, Occurrence, Effects, and Removal Methods. Chem Rev, 119 (6), 3510–3673.

Qin, L. T., Pang, X. R., Zeng, H. H., Liang, Y. P., Mo, L. Y., Wang, D. Q., & Dai, J. F. (2020). Ecological and human health risk of sulfonamides in surface water and groundwater of Huixian karst wetland in Guilin, China. Sci Total Environ, 708, 134552.

Rayner, C., & Munckhof, W. J. (2005). Antibiotics currently used in the treatment of infections caused by Staphylococcus aureus. Intern Med J, 35 *Suppl 2*, S3–16.

Reungoat, J., Escher, B. I., Macova, M., & Keller, J. (2011). Biofiltration of wastewater treatment plant effluent: effective removal of pharmaceuticals and personal care products and reduction of toxicity. Water Res, 45 (9), 2751–2762.

Rhouma, M., Fairbrother, J. M., Beaudry, F., & Letellier, A. (2017). Post weaning diarrhea in pigs: risk factors and non-colistin-based control strategies. Acta Veterinaria Scandinavica, 59 (1), 31.

Sachi, S., Ferdous, J., Sikder, M. H., & Azizul Karim Hussani, S. M. (2019). Antibiotic residues in milk: Past, present, and future. J Adv Vet Anim Res, 6 (3), 315–332.

Salama, N. A., Abou-Raya, S. H., Shalaby, A. R., Emam, W. H., & Mehaya, F. M. (2011). Incidence of tetracycline residues in chicken meat and liver retailed to consumers. Food Addit Contam Part B Surveill, 4 (2), 88–93.

Salaramoli, J., Heshmati, A., Kamkar, A., & Hassan, J. (2015). Effect of cooking procedures on tylosin residues in chicken meatball. Journal für Verbraucherschutz und Lebensmittelsicherheit, 11.

Schiller, C., Fröhlich, C. P., Giessmann, T., Siegmund, W., Mönnikes, H., Hosten, N., & Weitschies, W. (2005). Intestinal fluid volumes and transit of dosage forms as assessed by magnetic resonance imaging. Aliment Pharmacol Ther, 22 (10), 971–979.

Shalaby, A. R., Salama, N. A., Abou-Raya, S. H., Emam, W. H., & Mehaya, F. M. (2011). Validation of HPLC method for determination of tetracycline residues in chicken meat and liver. Food Chemistry, 124 (4), 1660–1666.

Shaltout, F. (2019). Impacts Of Different Types Of Cooking And Freezing On Antibiotic Residues In Chicken Meat. Food Science and Nutrition, 5.

Shirreffs, S. M., Watson, P., & Maughan, R. J. (2007). Milk as an effective post-exercise rehydration drink. Br J Nutr, 98 (1), 173–180.

Smith, B. I., Donovan, G. A., Risco, C., Littell, R., Young, C., Stanker, L. H., & Elliott, J. (1998). Comparison of various antibiotic treatments for cows diagnosed with toxic puerperal metritis. J Dairy Sci, 81 (6), 1555–1562.

Sobral, M. M. C., Cunha, S. C., Faria, M. A., & Ferreira, I. M. (2018). Domestic Cooking of Muscle Foods: Impact on Composition of Nutrients and Contaminants. Compr Rev Food Sci Food Saf, 17 (2), 309–333.

Spence, C. (2021). Explaining seasonal patterns of food consumption. International Journal of Gastronomy and Food Science, 24, 100332.

Stockwell, V. O., & Duffy, B. (2012). Use of antibiotics in plant agriculture. Rev Sci Tech, 31 (1), 199–210.

Thursby, E., & Juge, N. (2017). Introduction to the human gut microbiota. Biochem J, 474 (11), 1823–1836.

Tiseo, K., Huber, L., Gilbert, M., Robinson, T. P., & Van Boeckel, T. P. (2020). Global Trends in Antimicrobial Use in Food Animals from 2017 to 2030. Antibiotics (Basel), 9 (12).

Treiber, F. M., & Beranek-Knauer, H. (2021). Antimicrobial Residues in Food from Animal Origin-A Review of the Literature Focusing on Products Collected in Stores and Markets Worldwide. Antibiotics (Basel), 10 (5).

Ueland, Ø., Rødbotten, R., & Varela, P. (2022). Meat consumption and consumer attitudes – A Norwegian perspective. Meat Science, 192, 108920.

van Staa, T. P., Palin, V., Li, Y., Welfare, W., Felton, T. W., Dark, P., & Ashcroft, D. M. (2020). The effectiveness of frequent antibiotic use in reducing the risk of infection-related hospital admissions: results from two large population-based cohorts. BMC Medicine, 18 (1), 40.

Vanhaecke, T., Bretin, O., Poirel, M., & Tap, J. (2022). Drinking Water Source and Intake Are Associated with Distinct Gut Microbiota Signatures in US and UK Populations. J Nutr, 152 (1), 171–182.

Wang, Y., Uffelman, C. N., Bergia, R. E., Clark, C. M., Reed, J. B., Cross, T. L., Lindemann, S. R., Tang, M., & Campbell, W. W. (2023). Meat Consumption and Gut Microbiota: a Scoping Review of Literature and Systematic Review of Randomized Controlled Trials in Adults. Adv Nutr, 14 (2), 215–237.

Weill, F. X., Guesnier, F., Guibert, V., Timinouni, M., Demartin, M., Polomack, L., & Grimont, P. A. (2006). Multidrug resistance in Salmonella enterica serotype Typhimurium from humans in France (1993 to 2003). J Clin Microbiol, 44 (3), 700–708.

Xu, L., Zhang, H., Xiong, P., Zhu, Q., Liao, C., & Jiang, G. (2021). Occurrence, fate, and risk assessment of typical tetracycline antibiotics in the aquatic environment: A review. Sci Total Environ, 753, 141975.

Zhang, P. (2022). Influence of Foods and Nutrition on the Gut Microbiome and Implications for Intestinal Health. Int J Mol Sci, 23 (17).

