## Supplementary material for "Do You Know Your Daily Antibiotic Intake through Residues in Your Diet?": Table S1, Table S2, Table S3, Table S4, Table S5, Table S6, Figure S1, Figure S2

*Corresponding author:

**Table S1.** Linearity, limit of detection (LOD), and limit of quantification (LOQ) of each antibiotic’s calibration curve

| **Classes** | **Antibiotics** | **Linearity** | **LOD (μg/L)** | **LOQ (μg/L)** |
| --- | --- | --- | --- | --- |
| Tetracyclines | Tetracycline | 0.9995 | 10.949 | 33.179 |
|  | Oxytetracycline | 0.9997 | 8.498 | 25.750 |
| Penicillin | Amoxicillin | 0.9995 | 10.345 | 31.347 |
|  | Ampicillin | 0.9995 | 11.018 | 33.387 |
| Sulfonamides | Sulfadiazine | 0.9997 | 8.324 | 25.224 |
|  | Trimethoprim | 0.9993 | 12.534 | 37.980 |
| Macrolides | Erythromycin | 0.9999 | 5.751 | 17.428 |
|  | Tylosin | 0.9996 | 10.027 | 30.385 |
| Quinolones | Ciprofloxacin | 0.9996 | 8.933 | 27.071 |
|  | Enrofloxacin | 0.9994 | 11.707 | 35.475 |

**Figure S2.** The 10 target antibiotics (tetracycline, oxytetracycline, amoxicillin, ampicillin, sulfadiazine, trimethoprim, erythromycin, tylosin, ciprofloxacin, and enrofloxacin): reconstructed ion chromatogram for *m/z* 445, 461, 366, 350, 251, 291, 734, 916, 332, and 360, respectively.

Tetracycline

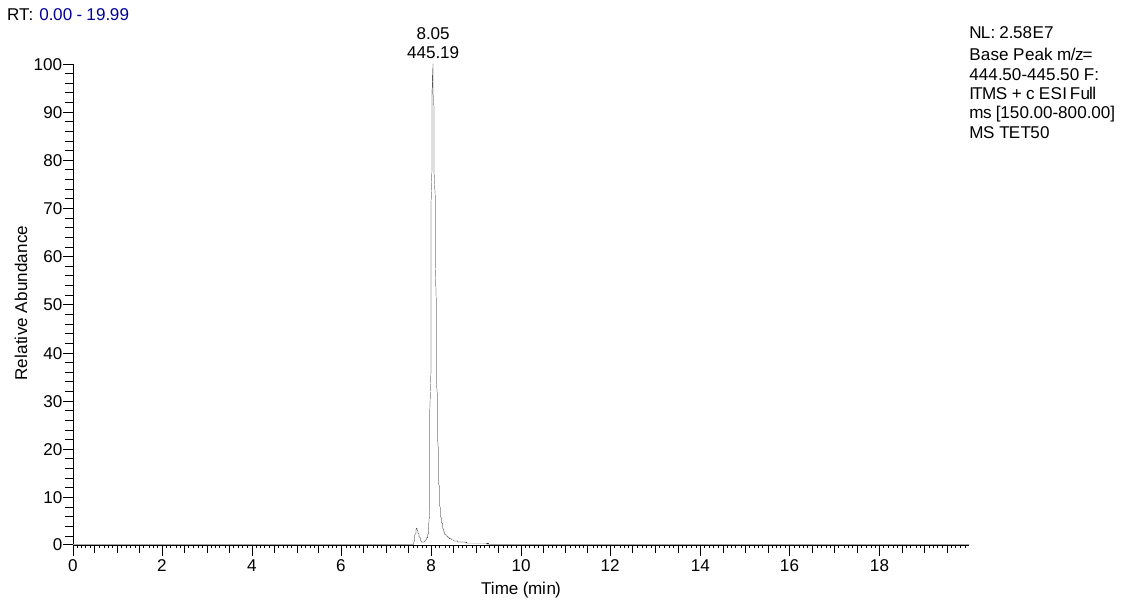

Oxytetracycline

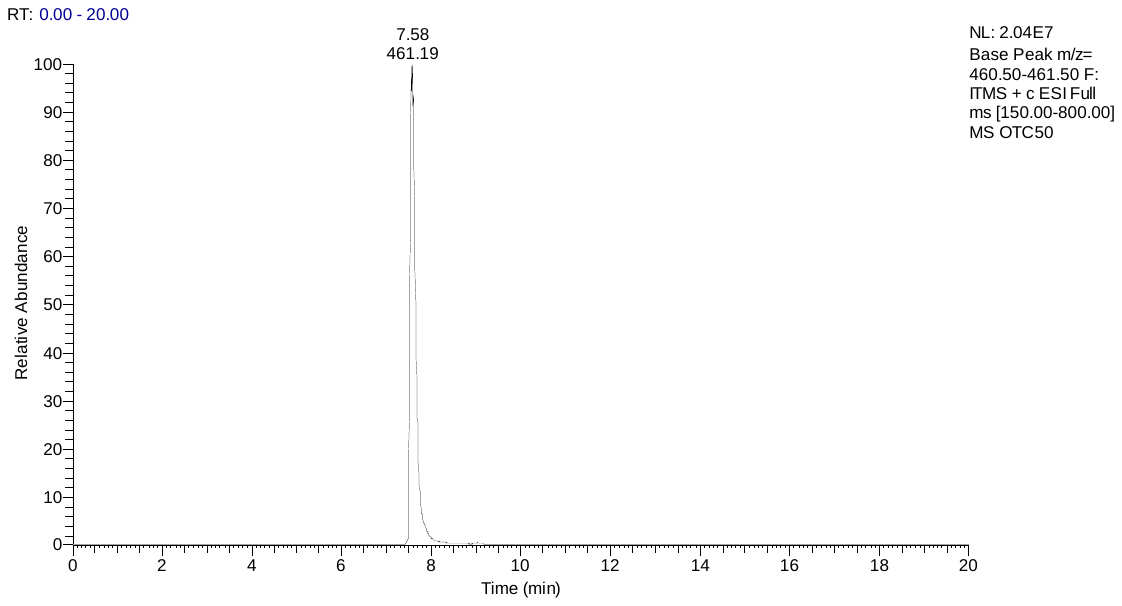

Amoxicillin

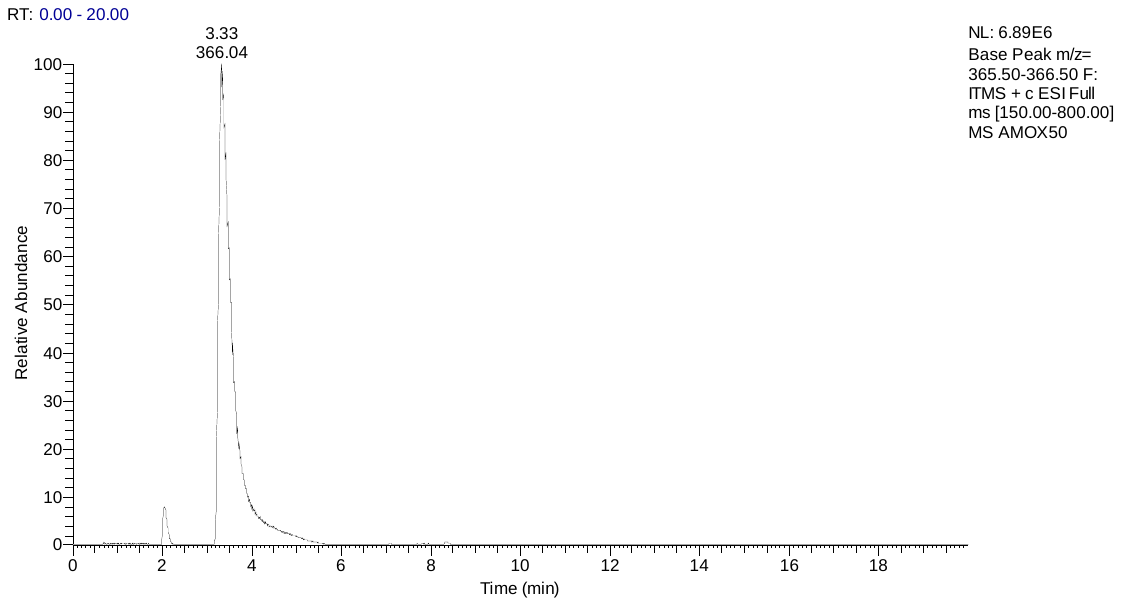

Ampicillin

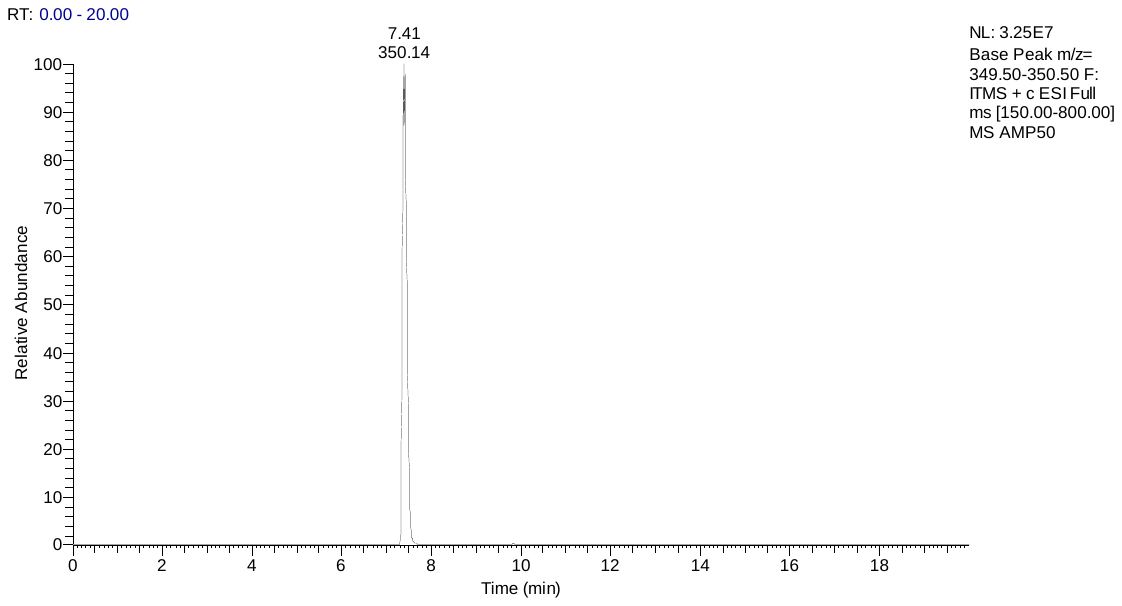

Sulfadiazine

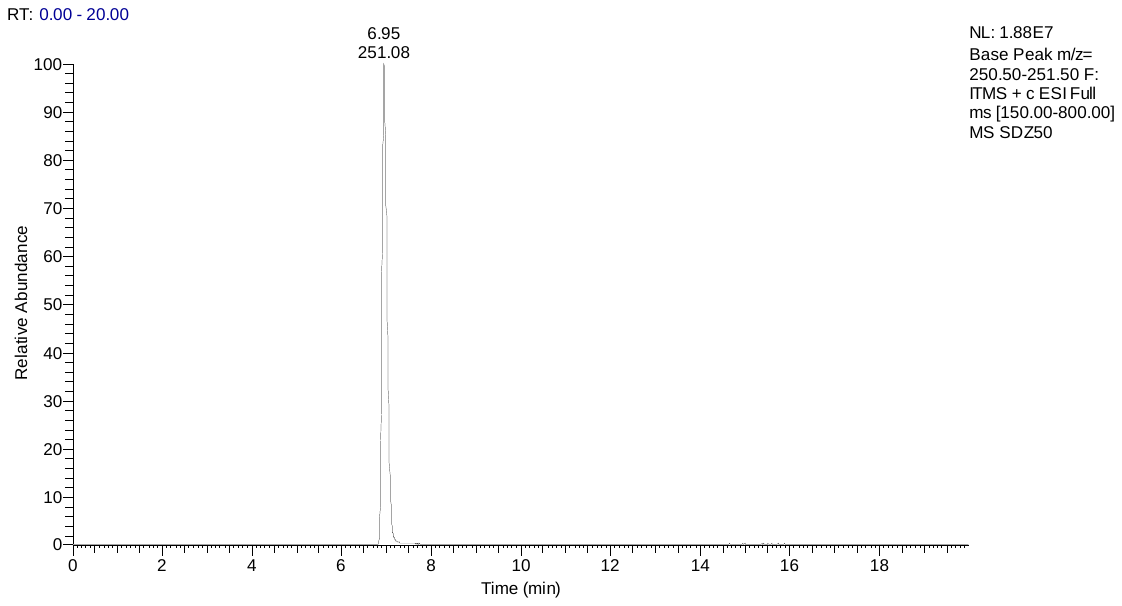

Trimethoprim

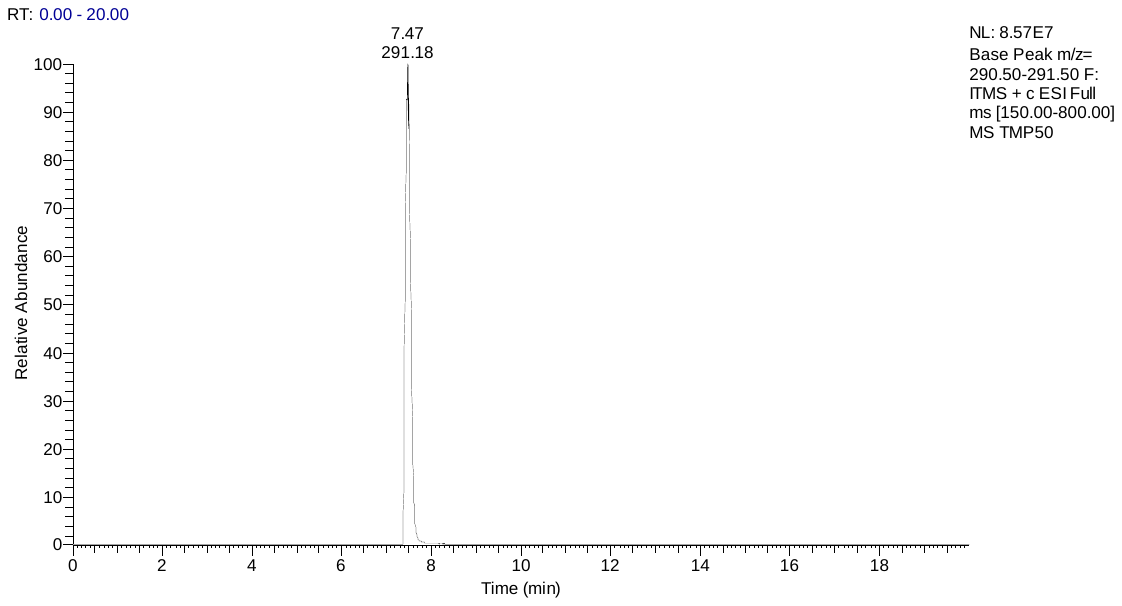

Erythromycin

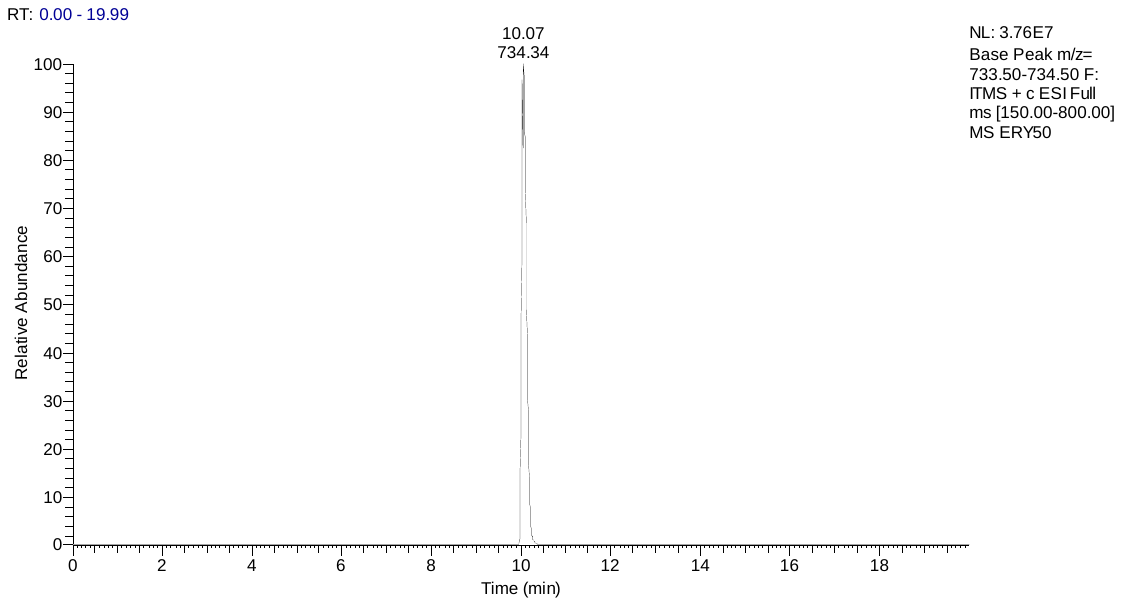

Tylosin

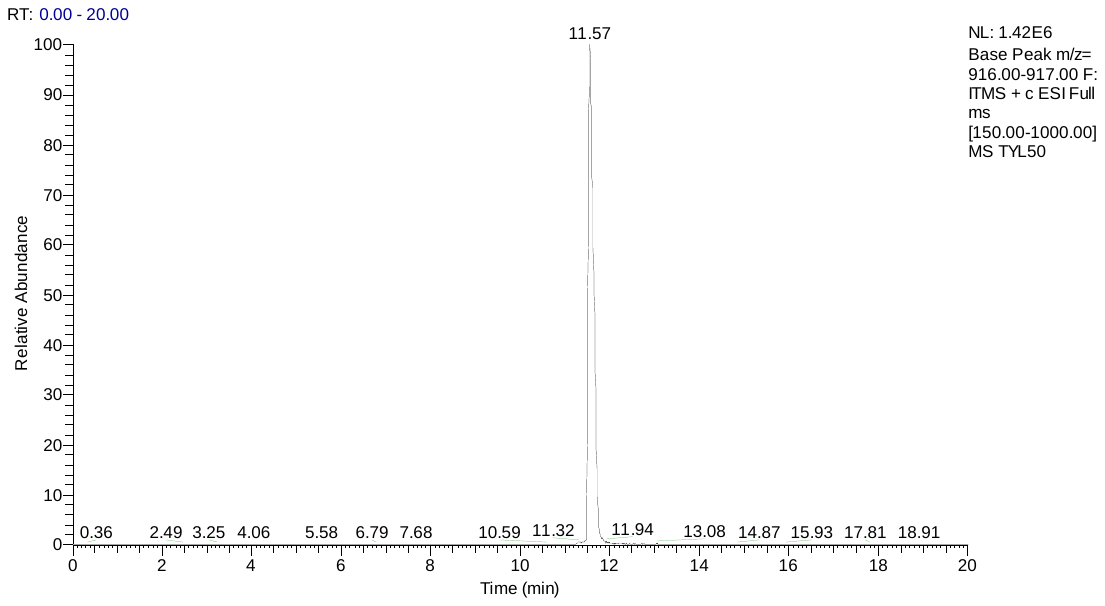

Ciprofloxacin

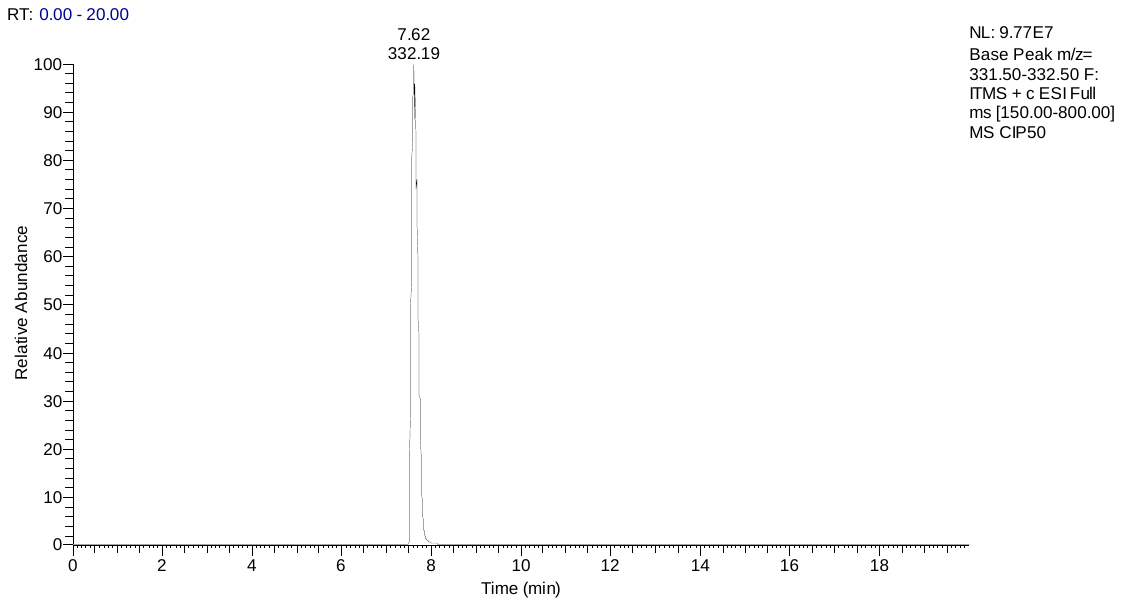

Enrofloxacin

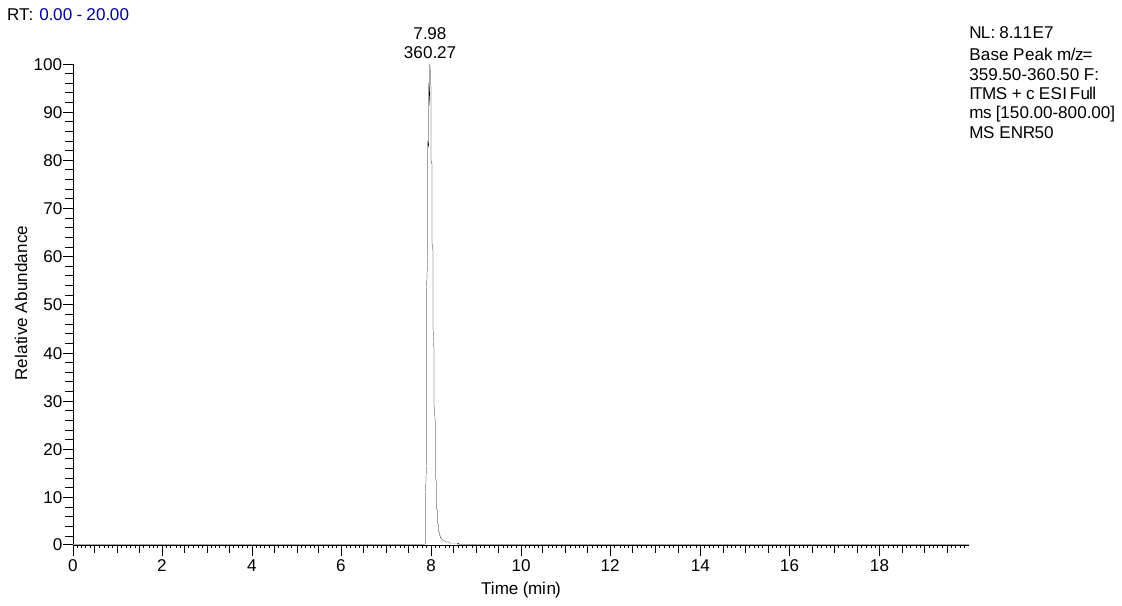

**Table S3.** The LTPE method validation using LC-MS analysis. Calibration curves and LTPE methods by parameters accuracy (%) and Relative Standard Deviation (RSD; %) using triplicates of nominal concentration (50, 100, and 500 μg/L) and measured concentration (μg/L).

| **Mean AB Standards Validation** | | | | | | | |
| --- | --- | --- | --- | --- | --- | --- | --- |
| **Antibiotic** | **MW** | **RT** | **Measured Area** | **Nominal concentration (μg/L)** | **Measured concentration (μg/L)** | **Accuracy (%)** | **RSD (%)** |
| Tetracycline | 445 | 8.13 | 195195 | 50.00 | 48.97 | 97.94 | 0.32 |
| Oxytetracycline | 461 | 7.73 | 166300 | 50.00 | 54.13 | 108.25 | 0.34 |
| Amoxicillin | 366 | 3.51 | 69293 | 50.00 | 52.72 | 105.44 | 0.51 |
| Ampicillin | 350 | 7.41 | 241966 | 50.00 | 55.61 | 111.22 | 0.20 |
| Sulfadiazine | 251 | 6.97 | 129273 | 50.00 | 48.67 | 97.34 | 0.44 |
| Trimethoprim | 291 | 7.53 | 846428 | 50.00 | 51.41 | 102.82 | 0.11 |
| Erythromycin | 734 | 10.04 | 226506 | 50.00 | 52.19 | 104.38 | 0.87 |
| Tylosin | 916 | 11.53 | 9916 | 50.00 | 50.54 | 101.08 | 0.37 |
| Ciprofloxacin | 332 | 7.76 | 418218 | 50.00 | 48.60 | 97.19 | 0.39 |
| Enrofloxacin | 360 | 8.06 | 704085 | 50.00 | 52.79 | 105.58 | 0.10 |

| **Mean AB Standards Validation** | | | | | | | |
| --- | --- | --- | --- | --- | --- | --- | --- |
| **Antibiotic** | **MW** | **RT** | **Measured Area** | **Nominal concentration (μg/L)** | **Measured concentration (μg/L)** | **Accuracy (%)** | **RSD (%)** |
| Tetracycline | 445 | 8.06 | 389463 | 100.00 | 101.92 | 101.92 | 0.04 |
| Oxytetracycline | 461 | 7.72 | 332537 | 100.00 | 104.75 | 104.75 | 0.08 |
| Amoxicillin | 366 | 3.52 | 138540 | 100.00 | 105.32 | 105.32 | 0.21 |
| Ampicillin | 350 | 7.46 | 482493 | 100.00 | 106.51 | 106.51 | 0.06 |
| Sulfadiazine | 251 | 6.91 | 257510 | 100.00 | 101.13 | 101.13 | 0.13 |
| Trimethoprim | 291 | 7.50 | 1693450 | 100.00 | 103.23 | 103.23 | 0.01 |
| Erythromycin | 734 | 10.04 | 445852 | 100.00 | 104.10 | 104.10 | 0.03 |
| Tylosin | 916 | 11.63 | 19256 | 100.00 | 102.15 | 102.15 | 0.76 |
| Ciprofloxacin | 332 | 7.76 | 841564 | 100.00 | 101.33 | 101.33 | 0.02 |
| Enrofloxacin | 360 | 8.05 | 1408338 | 100.00 | 102.26 | 102.26 | 0.02 |

| **Mean AB Standards Validation** | | | | | | | |
| --- | --- | --- | --- | --- | --- | --- | --- |
| **Antibiotic** | **MW** | **RT** | **Measured Area** | **Nominal concentration (μg/L)** | **Measured concentration (μg/L)** | **Accuracy (%)** | **RSD (%)** |
| Tetracycline | 445 | 8.07 | 1923085 | 500.00 | 519.42 | 103.88 | 0.60 |
| Oxytetracycline | 461 | 7.64 | 1660534 | 500.00 | 509.00 | 101.80 | 0.34 |
| Amoxicillin | 366 | 3.62 | 690274 | 500.00 | 522.92 | 104.58 | 0.48 |
| Ampicillin | 350 | 7.45 | 2419374 | 500.00 | 515.28 | 103.06 | 0.51 |
| Sulfadiazine | 251 | 6.92 | 1289961 | 500.00 | 523.26 | 104.65 | 0.42 |
| Trimethoprim | 291 | 7.52 | 8476638 | 500.00 | 518.45 | 103.69 | 0.12 |
| Erythromycin | 734 | 10.01 | 2221920 | 500.00 | 524.26 | 104.85 | 0.28 |
| Tylosin | 916 | 11.68 | 91891 | 500.00 | 506.37 | 101.27 | 0.92 |
| Ciprofloxacin | 332 | 7.83 | 4182029 | 500.00 | 517.20 | 103.44 | 0.43 |
| Enrofloxacin | 360 | 8.08 | 7036687 | 500.00 | 497.68 | 99.54 | 0.03 |

| **Mean AB LTPE Validation** | | | | | | | |
| --- | --- | --- | --- | --- | --- | --- | --- |
| **Antibiotic** | **MW** | **RT** | **Measured Area** | **Nominal concentration (μg/L)** | **Measured concentration (μg/L)** | **Accuracy (%)** | **RSD (%)** |
| Tetracycline | 445 | 7.97 | 340633 | 100.00 | 88.61 | 88.61 | 0.31 |
| Oxytetracycline | 461 | 7.65 | 295535 | 100.00 | 93.48 | 93.48 | 0.03 |
| Amoxicillin | 366 | 1.94 | 117015 | 100.00 | 88.97 | 88.97 | 0.68 |
| Ampicillin | 350 | 7.39 | 414491 | 100.00 | 92.12 | 92.12 | 0.76 |
| Sulfadiazine | 251 | 6.91 | 236253 | 100.00 | 92.44 | 92.44 | 0.77 |
| Trimethoprim | 291 | 7.43 | 1515047 | 100.00 | 92.32 | 92.32 | 0.63 |
| Erythromycin | 734 | 10.05 | 397927 | 100.00 | 92.76 | 92.76 | 0.75 |
| Tylosin | 772 | 11.55 | 17439 | 100.00 | 92.12 | 92.12 | 0.46 |
| Ciprofloxacin | 332 | 7.68 | 772549 | 100.00 | 92.73 | 92.73 | 0.60 |
| Enrofloxacin | 360 | 7.97 | 1199189 | 100.00 | 87.57 | 87.57 | 0.18 |

**Table S4**. The recovery of LTPE method using 100 µg/L of 10 antibiotic mixture stock solution, and the recovery of using triplicates of pork chop matrix spiked with a 100 μg/L of 10 antibiotics mixture.

| **Mean Spiked Meat Validation** | | | | | | | |
| --- | --- | --- | --- | --- | --- | --- | --- |
| **Antibiotic** | **RT** | **Measured Area** | **AB standard nominal concentration (μg/L)** | **Total AB concentration from spiked meat (μg/kg)** | **AB concentration from non-spiked meat (μg/kg)** | **Accuracy of spiked meat (%)** | **Accuracy of AB LTPE (%)** |
| Tetracycline | 8.06 | 345155 | 100.00 | 89.84 |  | 89.84 | 88.61 |
| Oxytetracycline | 7.62 | 303293 | 100.00 | 95.84 |  | 95.84 | 93.48 |
| Amoxicillin | 2.05 | 2249514 | 100.00 | 1708.64 | 1616.09 | 92.55 | 88.97 |
| Ampicillin | 7.48 | 414539 | 100.00 | 92.13 |  | 92.13 | 92.12 |
| Sulfadiazine | 6.81 | 523468 | 100.00 | 209.94 | 116.19 | 93.75 | 92.44 |
| Trimethoprim | 7.68 | 9084446 | 100.00 | 555.45 | 461.68 | 93.77 | 92.32 |
| Erythromycin | 10.37 | 402559 | 100.00 | 93.85 |  | 93.85 | 92.76 |
| Tylosin | 11.58 | 17858 | 100.00 | 94.43 |  | 94.43 | 92.12 |
| Ciprofloxacin | 7.87 | 785378 | 100.00 | 94.33 |  | 94.33 | 92.73 |
| Enrofloxacin | 8.06 | 1228007 | 100.00 | 89.60 |  | 89.60 | 87.57 |

**Table S5.** Minimum, maximum and mean consumption (g or mL) on dairy product consumption.

| **Time** | **Summer (g or mL)** | | | | | | **Winter (g or mL)** | | | | | |
| --- | --- | --- | --- | --- | --- | --- | --- | --- | --- | --- | --- | --- |
|  | **Day 1** | | | **Day 2** | | | **Day 1** | | | **Day 2** | | |
|  | **Min.** | **Max.** | **Mean** | **Min.** | **Max.** | **Mean** | **Min.** | **Max.** | **Mean** | **Min.** | **Max.** | **Mean** |
| 0600-0659 | - | - | - | - | - | - | - | - | - | - | - | - |
| 0700-0759 | 50 | 250 | 88 | 50 | 50 | 50 | 50 | 250 | 88 | 50 | 50 | 50 |
| 0800-0859 | 50 | 350 | 130 | 50 | 350 | 131 | 50 | 300 | 122 | 50 | 300 | 142 |
| 0900-0959 | 150 | 150 | 150 | - | - | - | 150 | 150 | 150 | - | - | - |
| 1000-1059 | - | - | - | 100 | 100 | 100 | - | - | - | 100 | 100 | 100 |
| 1100-1159 | - | - | - | - | - | - | - | - | - | - | - | - |
| 1200-1259 | - | - | - | - | - | - | - | - | - | - | - | - |
| 1300-1359 | 50 | 108 | 69 | 50 | 50 | 50 | 50 | 100 | 67 | 50 | 100 | 67 |
| 1400-1459 | - | - | - | 50 | 50 | 50 | 100 | 180 | - | 50 | 100 | 75 |
| 1500-1559 | - | - | - | - | - | - | 180 | 180 | - | - | - | - |
| 1600-1659 | 100 | 100 | 100 | 100 | 100 | 100 | 100 | 100 | 100 | 100 | 100 | 100 |
| 1700-1759 | - | - | - | 100 | 100 | 100 | - | - | - | 100 | 100 | 100 |
| 1800-1859 | 65 | 65 | 65 | 65 | 65 | 65 | 65 | 180 | 123 | 65 | 65 | 65 |
| 1900-1959 | - | - | - | 50 | 50 | 50 | - | - | - | 50 | 50 | 50 |
| 2000-2059 | 180 | 180 | 180 | - | - | - | 180 | 180 | 180 | 40 | 300 | - |
| 2100-2159 | - | - | - | - | - | - | - | - | - | - | - | - |
| 2200-2259 | - | - | - | - | - | - | - | - | - | - | - | - |
| 2300-2359 | - | - | - | - | - | - | - | - | - | - | - | - |

**Table S6**. Minimum, maximum, median, and mean consumption (g or mL) on water consumption.

| **Time** | **Summer (mL)** | | | | | | | | **Winter (mL)** | | | | | | | |
| --- | --- | --- | --- | --- | --- | --- | --- | --- | --- | --- | --- | --- | --- | --- | --- | --- |
|  | **Day 1** | | | | **Day 2** | | | | **Day 1** | | | | **Day 2** | | | |
|  | **Min.** | **Max.** | **Median** | **Mean** | **Min.** | **Max.** | **Median** | **Mean** | **Min.** | **Max.** | **Median** | **Mean** | **Min.** | **Max.** | **Median** | **Mean** |
| 0700-0759 | 350 | 350 | 350 | 350 | 200 | 350 | 275 | 275 | 120 | 350 | 150 | 207 | 200 | 300 | 200 | 233 |
| 0800-0859 | 180 | 420 | 200 | 241 | 60 | 400 | 250 | 252 | 180 | 500 | 320 | 322 | 170 | 500 | 300 | 315 |
| 0900-0959 | 60 | 700 | 225 | 278 | 40 | 500 | 325 | 282 | 150 | 500 | 225 | 268 | 180 | 500 | 250 | 304 |
| 1000-1059 | 60 | 350 | 200 | 202 | 60 | 350 | 225 | 210 | 40 | 350 | 120 | 140 | 140 | 350 | 215 | 219 |
| 1100-1159 | 180 | 250 | 200 | 210 | 60 | 180 | 150 | 130 | 80 | 250 | 140 | 140 | 60 | 300 | 150 | 158 |
| 1200-1259 | 120 | 500 | 180 | 243 | 180 | 500 | 250 | 306 | 40 | 400 | 135 | 164 | 180 | 600 | 325 | 408 |
| 1300-1359 | 150 | 1140 | 250 | 368 | 180 | 850 | 300 | 374 | 140 | 700 | 300 | 354 | 180 | 1000 | 375 | 453 |
| 1400-1459 | 150 | 700 | 350 | 376 | 120 | 500 | 200 | 250 | 100 | 400 | 200 | 219 | 120 | 300 | 180 | 201 |
| 1500-1559 | 150 | 500 | 250 | 303 | 60 | 350 | 180 | 242 | 50 | 400 | 150 | 166 | 180 | 500 | 300 | 310 |
| 1600-1659 | 180 | 750 | 465 | 465 | 60 | 500 | 180 | 247 | 100 | 400 | 200 | 244 | 60 | 200 | 100 | 126 |
| 1700-1759 | 40 | 500 | 300 | 285 | 180 | 350 | 275 | 270 | 100 | 400 | 300 | 275 | 180 | 350 | 300 | 276 |
| 1800-1859 | 180 | 500 | 225 | 268 | 180 | 300 | 250 | 254 | 80 | 500 | 190 | 213 | 100 | 500 | 250 | 254 |
| 1900-1959 | - | 1050 | 360 | 398 | 180 | 1050 | 250 | 384 | 180 | 750 | 200 | 339 | 180 | 800 | 275 | 326 |
| 2000-2059 | 150 | 700 | 190 | 263 | 180 | 500 | 200 | 288 | 100 | 500 | 190 | 260 | 100 | 360 | 200 | 203 |
| 2100-2159 | 180 | 360 | 200 | 235 | 150 | 360 | 200 | 228 | 180 | 200 | 200 | 193 | 100 | 300 | 180 | 194 |
| 2200-2259 | 180 | 250 | 215 | 215 | 180 | 180 | 180 | 180 | 120 | 180 | 180 | 160 | 180 | 200 | 190 | 190 |
| 2300-2359 | 200 | 200 | 200 | 200 | 180 | 200 | 250 | 227 | 150 | 200 | 175 | 175 | 170 | 180 | 180 | 177 |
